# ESTIMATING THE INCIDENCE OF SARS-COV-2 INFECTIONS IN 2020 IN BELGIUM BY JOINTLY MODELLING SEROPREVALENCE, HOSPITALIZATION AND MORTALITY DATA

**DOI:** 10.1101/2025.07.02.25330709

**Authors:** Toon Braeye, Robby De Pauw, Laurence Geebelen, Steven Abrams, Isabelle Desombere, Niel Hens, Naïma Hammami, Mathieu Roelants, Sereina A. Herzog

## Abstract

**BACKGROUND:** The incidence of SARS-CoV-2 infections was not directly observed during the pandemic. We estimated incidences and the associated burden, through the infection fatality rate (IFR) and infection hospitalization rate (IHR), by jointly modeling seroprevalence, hospitalization, and mortality data.

**METHODS:** We used a hierarchical Bayesian model to fit a spline, representing incidence, to the observed number of hospitalizations, deaths and seroprevalence results. Two cross-sectional seroprevalence studies on the prevalence of SARS-CoV-2 antibodies in the general population collected 37,235 samples in the age interval 18 to 74 years across 30 time points between March 2020 and January 2021. One study analyzed residual laboratory samples with the Euroimmun Anti-SARS-CoV-2 ELISA. The other study analyzed blood donor samples with the Wantai Ab ELISA. Data on the sensitivity and specificity of the serological tests were obtained from published research. Time-varying IFR and IHR estimates and their associated delay distributions were estimated within the same model.

**RESULTS:** By the end of January 2021, we estimated 19% (95%CrI 18%-21%), 13% (95%CrI 12%-15%) and 11% (95%CrI 8%-13%) of the Belgian 18-49, 50-64 and 65-74 year-olds to have been infected with SARS-CoV-2. Infections occurred in two large waves with few infections in between. The first wave mostly affected the younger age group, with a peak weekly incidence of 2.0% (95%CrI 1.7%-2.3%) end of March 2020, while weekly incidences were more comparable during the second wave end of October 2020: 1.6% (95%CrI 1.2%-2%) for 65-74 year-olds and 2.8% (95%CrI 2.3%-3.3%) for 18-49 year-olds.

Both the hospitalization and fatality rates declined over time. Among persons aged 65 to 74 years the hospitalization rate declined from 9.9% (95%CrI 7.2-13.2) to 5.1% (95%CrI 3.2-6.3) and fatality rates from 2.8% (95%CrI 2.0%-3.8%) to 1.3% (95%CrI 0.9%-1.7%). IHR and IFR were considerably lower in the younger age groups.

**CONCLUSION:** During 2020, an estimated 16.2% (95%CrI 15.1%-17.3%) of the Belgian adult population was infected with SARS-CoV-2. These infections occurred primarily in two waves, one in March and another one in October 2020, with minimal transmission in between. Severe disease was more prevalent among older age groups and earlier on in 2020.

## 2. Introduction

In Belgium, the first infections with SARS-CoV-2 were reported by the end of February 2020. A rapid increase in cases resulted in a strict lockdown on the 18^th^ of March 2020 and a first peak in cases was reached at the beginning of April [1]. During this first lockdown no social interactions were allowed outside of the household and non-essential travel was forbidden. The gradual ease of restrictions started in May 2020, but many measures remained in place during the summer of 2020. Because of a resurgence of cases by the end of September, a new lockdown was implemented in mid-October. The second lockdown involved less restrictions, allowing for, for example, a single contact outside of the household. While case counts, COVID-19 related hospitalizations and deaths during 2020 have been well documented [2], the actual number of infections remains unknown. The case count can only be considered as a poor proxy since limited testing capacity and a restrictive initial case definition defined the testing policy in the first half of 2020 [3]. In addition, a proportion of SARS-CoV-2 infections will remain asymptomatic and adherence to testing policy will be incomplete further limiting the detection of infections.

Since the majority of infected individuals produce antibodies to SARS-CoV-2 that persist for at least several months [4–7], serological screening can be used to estimate the proportion of the population previously infected with SARS-CoV-2 [8,9]. While seroprevalence studies were very common during the COVID-19 pandemic, especially in the pre-vaccination era, there is no consensus on how best to interpret their results [10]. Qualitative seroprevalence results indicate whether an antibody titer is detectable. Titers however require time to build-up after infection and will typically decline over longer time periods. In addition to time-varying titers, serological tests are imperfect. Some researchers have explicitly selected serological tests with stable longitudinal test performance or combined results from different immunoassays to improve sensitivity and specificity [11,12]. Alternatively, studies used Rogan-Gladen type estimators to correct for incomplete sensitivity and specificity [13]. Researchers might however find that different specificity and especially sensitivity estimates exist for a serological test and that the outcome of their work is sensitive to these estimates. For example, different estimates have resulted in considerable discrepancies in infection (fatality) rates [11,14]. In previous work we have presented a framework that allows for the estimation of test-specific sensitivity given time since infection, age and clinical severity of the infection. The suggested distribution captured the process of antibody build-up to detectable levels, seroconversion, and subsequent waning of antibodies, seroreversion, using a Weibull-Bi-exponential distribution. Comparable approaches have been discussed in other studies on modelling of seroprevalence results [15,16].

In Belgium two prospective cross-sectional seroprevalence studies were set up to estimate the level of past SARS-CoV-2 infection. One study collected residual laboratory samples (RLS) [17] while the other collected blood donors samples (BDS) [18]. Both studies aimed to sample from the general population, but differences in the study’s set-up, cohorts, timing and tests used complicate the direct combination of these data sources. For example, the qualitative serological tests used: the EuroImmun IgG (used by RLS) and Wantai total Ab (used by BDS), have different reported diagnostic characteristics. The EuroImmun test’s sensitivity estimates vary as widely as from 64.1% to 94.4% [19], with a decline in test sensitivity with time since infection [11,20,21]. The Wantai test has a reported 94.2-98.0% sensitivity and no reported seroreversion for at least 13-15 months after infection [22–26]. A statistical framework is necessary to account for these differences.

We aim to estimate the incidence of SARS-CoV-2 infections and the associated infection fatality (IFR) and hospitalization (IHR) rates for the Belgian general population aged 18 to 74 years old from two prospective cross-sectional seroprevalence studies and surveillance data on COVID-19 related hospitalizations and deaths.

## 3. Methods

We first present the seroprevalence and surveillance data. We then present the model.

### 3.1. DATA SOURCES

#### 3.1.1. Seroprevalence

Both seroprevalence studies followed a repeated cross-sectional design and collected the first samples on 30 March 2020. The last collection round for the RLS was on 12 October 2020. After each collection round a report was created and seroprevalence results were presented online. Only data on age (in years), sex and region of residence were collected alongside the sample. Researchers could not identify participants. The last collection round considered here for the BDS included samples collected at the end of January 2021. The mass rollout of vaccination in Belgium started in January 2021 and prioritized nursing home residents and staff. The impact of vaccination on antibody titers in the general population before February 2021 is therefore expected to be limited. An earlier end date would not allow us to include the decreasing phase of the second surge of COVID-19 in Belgium. We included only persons between the ages of 18 and 74 years.

The RLS was coordinated by the Centre for the Evaluation of Vaccination, Vaccine & Infectious Disease Institute, at the University of Antwerp. A total of 16,547 serum samples were included from 7 collection rounds with approximately one-month intervals between consecutive rounds. Samples were collected from ambulatory patients through 10 private diagnostic laboratories and analyzed using the Anti-SARS-CoV-2 IgG ELISA from Euroimmun (cat n° EI 2606-9601G; Medizinische Labordiagostika AG, Germany). This semi-quantitative test detects IgG antibodies against the SARS-CoV-2 S1 protein. Details and results from this study have been published in Herzorg et al. [17]. The RLS excluded samples received from COVID-19 specific hospital units and triage centers.

The BDS was coordinated by Sciensano [18], Belgium’s institute for public health, in cooperation with the red cross. A total of 20,688 plasma samples were included from 23 collection rounds with 14-day intervals. The Wantai SARS-CoV-2 Ab ELISA (cat n° WS-1096; Beijing Wantai Biological Pharmacy Enterprice Co. Ltd., China) was used for the qualitative detection of total antibodies directed to the Receptor-Binding Domain (RBD). This ELISA detects anti-RBD IgG, IgA and IgM concomitantly.

Exclusion criteria for donation were being underage, severe disease or anemia, persons with a previously confirmed or possible COVID-19 infection and having had a close contact with a confirmed SARS-CoV-2 case in the last 14 days. At each time point, a sample was drawn, out of all blood donations, of 425 subjects for the Flemish Region, 320 for the Walloon Region and 155 for the Brussels Capital Region following the marginal distribution of sex, age and province within each region.

#### 3.1.2. Data on COVID-19 mortality and hospitalization

Belgian COVID-19 mortality surveillance has been described elsewhere [27]. In short, the surveillance aimed to be exhaustive by including data from multiple sources and by including both laboratory-confirmed and clinically suspect deaths of COVID-19. The number of deaths within the surveillance data compare closely to all-cause excess mortality [1] and ‘cause of death’-certificates [28]. We excluded deaths in nursing homes as these occurred outside the general population.

A Clinical Hospital Survey (CHS) collected data on all COVID-19 cases within the hospital [29]. We excluded asymptomatic cases found during screening within the hospital. There is no formal assessment of the coverage of the survey in 2020, but in comparison to an exhaustive survey on hospital intakes and capacity [29] it appeared to be close to 100% (S1 Fig).

### 3.2. THE MODEL

We estimated weekly incidences of SARS-CoV-2 infections and jointly fitted these to seroprevalence, hospital and mortality data. A hierarchical Bayesian model combined these three inputs: (1) the seroprevalence studies, RLS and BDS, given test and ‘time-since-infection’ specific sensitivity and specificity, (2) the number of hospitalizations for COVID-19 given ‘calendar time’-varying IHR and (3) the number of COVID-19 deaths given ‘calendar time’-varying IFR.

#### 3.2.1. Observed seropositivity

We fitted the number of observed positive samples to the total number of samples collected at week (𝑡) by age group (18-49, 50-64, 65-74 years) and by test (EuroImmun or Wantai). To account for the expected additional variance associated with seroprevalence samples of convenience we used a beta-binomial likelihood [30]. Specifically, we modeled the probability of detectable antibodies (coined seropositivity) 𝑃𝑜𝑏𝑠_𝑡𝑒𝑠𝑡,𝑎𝑔𝑒,𝑡_ as informing a beta-distributed random probability 𝑃𝑠𝑎𝑚𝑝𝑙𝑒_𝑡𝑒𝑠𝑡,𝑎𝑔𝑒,𝑡_, which was then used as the parameter in the binomial likelihood for the observed positive samples.

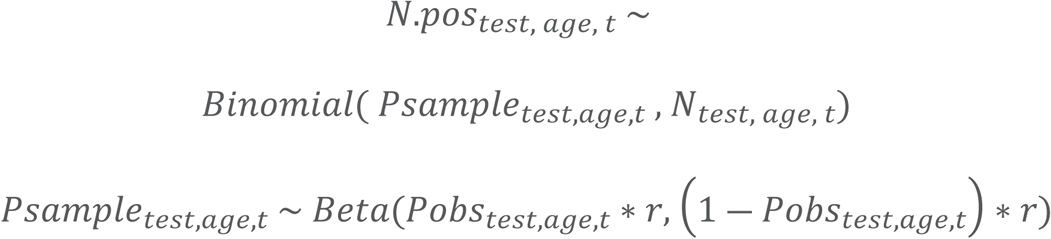

The overdispersion parameter r was given an informative gamma prior (shape=5, scale=1000) so by default the overdispersion should be limited. The prior however has sufficient variance to include overdispersion if necessary. We define the cumulative incidence (𝑐𝑢𝑚𝑢𝑙𝑎𝑡𝑖𝑣𝑒.𝑖𝑛𝑐𝑖𝑑𝑒𝑛𝑐𝑒_𝑡_) as the cumulative proportion of individuals that were infected with SARS-CoV-2 in the population. We obtained it by summing weekly incidences. The cumulative incidence differs from the seroprevalence because seroprevalence will include false positive (persons not infected, but with positive serological test results) and false negatives (persons previously infected without seroconversion or already seroreverted). To obtain the seroprevalence by week, test and age group (𝑃𝑜𝑏𝑠_𝑡𝑒𝑠𝑡,𝑎𝑔𝑒,𝑡_) for the incidence up to week 𝑡, we used a Rogan-Gladen type estimator [13,31].

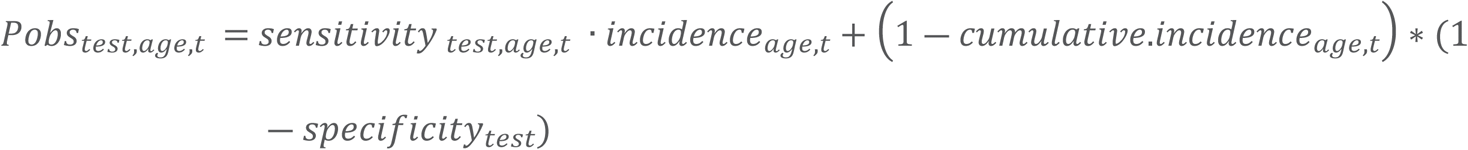

𝑠𝑒𝑛𝑠𝑖𝑡𝑖𝑣𝑖𝑡𝑦 _𝑡𝑒𝑠𝑡,𝑎𝑔𝑒,𝑡_ · 𝑖𝑛𝑐𝑖𝑑𝑒𝑛𝑐𝑒_𝑎𝑔𝑒,𝑡_ represents the cumulative dot product of the age-specific time-varying test sensitivity, accounting for the difference between the week of infection and the week of collection for the serosurvey, and the incidence at week 𝑡.

#### 3.2.2. Sensitivity

Informative prior distributions for the time-varying, test and age-specific sensitivity were based on accompanying work (*paper in submission*). In short, we combined data from published studies and Belgium’s SARS-CoV-2 laboratory testing to obtain scaled Weibull-Bi-exponential distributions for the sensitivity of the EuroImmun and Wantai test. Normal approximations of the posterior distributions of the shape, scale and exponential parameters associated with the distributions for seroconversion and seroreversion processes served as informative priors in the current model. Since our previous work accounted for infection severity, we could address a key limitation of both the BDS and RLS datasets: their exclusion of hospitalized cases. Consequently, our informative priors only covered asymptomatic and symptomatic infections that did not require hospitalization. Distributions representing the time-varying sensitivity are presented in Fig 2.

We included a sensitivity analysis to investigate the effect of having a single estimate for the tests’ sensitivity. These sensitivity estimates, one per test, are constant over the time since infection, but can be regarded as partially accounting for antibody decay as they have been derived from data obtained at different time points [32]. Sensitivity of the EuroImmune test is included as in Herzog et al. [17] (154 true positive and 27 false negative, approximately 85% sensitivity). For the Wantai test, sensitivity is included based on Harritshøj et al. [33] (145 true positive and 5 false negative, approximately 97% sensitivity). Results are presented in the S6 Fig.

#### 3.2.3. Specificity

The specificity included in the Rogan-Gladen estimator was estimated from published research on samples collected prior to 2020. We included five studies for both the EuroImmun [33–37] (1834 true negative and 22 false positives, approximately 98.8% specificity) and Wantai [33,35–38] test (2066 true negatives and 20 false positives, approximately 99% specificity). A breakdown of the data used per study is presented in the supporting information. Specificity represents the probability for obtaining negative samples from non-infected individuals and it was included as a binomial likelihood function in the model. A beta-prior (shape=20, scale=0.1) was chosen for the specificity parameter.

#### 3.2.4. Proportion infected

The incidence at week 𝑡 (𝑖𝑛𝑐𝑖𝑑𝑒𝑛𝑐𝑒_𝑎𝑔𝑒,𝑡_) was modelled as a function of time using a cubic B-splines without penalization with 10 internal knots. A spline-based approach allows for a flexible model that describes the infection process in the population. The spline coefficients were given weakly informative normal priors with mean 0 and standard deviation 100.

#### 3.2.5. Delay distributions: Mortality and Hospitalization

Binomial likelihoods were used to fit the number of hospitalizations and deaths to the numbers of infected individuals. The IHR and IFR were used as the probability parameter of the distribution. The number of infected individuals was obtained by multiplying the incidence with the population numbers.

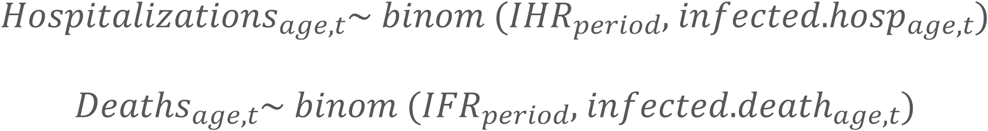

Discrete delay distributions were used to estimate the number of infections relevant for the number of hospitalizations or deaths at week 𝑡. For both hospitalizations and deaths, delays could amount to a maximum of eight weeks.

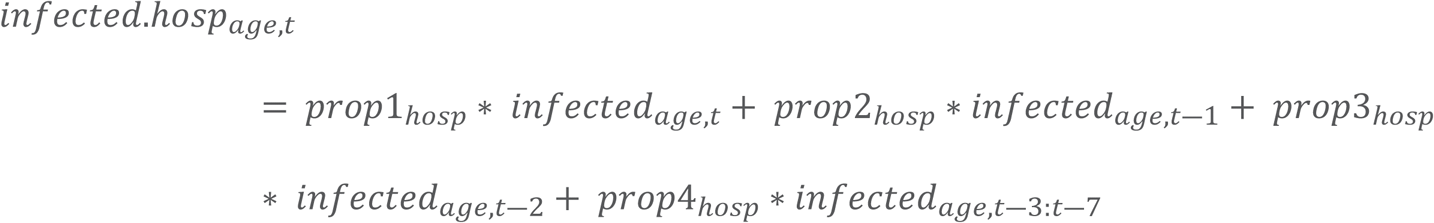

Given the Dirichlet prior distribution for the proportions (𝑝𝑟𝑜𝑝) in week 𝑡 ― 1:8, proportions over the delay period summed to one.

We allowed the IHR and IFR to vary over pre-defined periods. Hospital intake policy and healthcare seeking behavior varied over time as it depended, amongst other, on hospital capacity. To accommodate to this variation, six periods were introduced: first lockdown (study start – 9 May), first interwave period (10 May to 18 July), summer surge (19 July to 22 August), second interwave period (23 August to 03 October), second lockdown (4 October to 12 December) and finally the third interwave period (13 December 2020 to 31 January 2021). Both IHR and IFR had uniform priors on [0,1].

### 3.3. ETHICS

The BDS protocol was approved by the ethical committee of UZ Leuven (ethical approval S63932-UZ Leuven). Donors were excluded from the study if no consent was given for the use of residual blood for scientific research.

The RLS protocol was approved by the Ethics Committee of the University Hospital Antwerp-University of Antwerp on 30 March 2020 (ref 20/13/158; Belgian Number B3002020000047). The committee agreed with inclusion without informed consent, on the condition of the samples being collected anonymously.

### 3.4. SOFTWARE AND CODE

Hierarchical Bayesian models were fitted using the Nimble package in R [39]. Code is provided in the supporting information. This package uses Markov Chain Monte Carlo (MCMC) sampling to sample from posterior distributions. We used three MCMC chains with 10,000 iterations each and a burn-in of 4,000 iterations. Convergence of the different chains was checked using the Gelman-Rubin statistic.

## 4. Results

### 4.1. DATA DESCRIPTION

Both the BDS and RLS collected samples for six months in 2020. This led to a combined number of samples of around 4,100 per month. For March 2020, the BDS only sampled the Flemish region, leading to a lower total number of samples for March (N = 3,349). For the summer period and after October only the BDS collected samples (N = 1,799-3,035 per collection round). The combined unweighted seroprevalence increased over time to reach 19% in the youngest age group (18-49 years) and 13% in the oldest age group (65-74 years) by the end of January 2021. After weighting the data to correct for unrepresentative sampling from regional age and sex distributions, the seroprevalence for the youngest age group decreased to 18%, while it remained at 13% for the oldest age group (Table 1).

**Table 1:** Number of samples and the proportion of positives (unweighted) by collection month per age group (A) and per study (B), March 2020-January 2021, Belgium.

(A) Numbers per age group
| Age Group | 2020-03 | 2020-04 | 2020-05 | 2020-06 | 2020-07 | 2020-08 | 2020-09 | 2020-10 | 2020-11 | 2020-12 | 2021-01 |
| --- | --- | --- | --- | --- | --- | --- | --- | --- | --- | --- | --- |
| Number of samples |  |  |  |  |  |  |  |  |  |  |  |
| 18-49 | 1861 | 2420 | 2506 | 3679 | 1096 | 1622 | 2416 | 2386 | 1085 | 1093 | 1777 |
| 50-64 | 904 | 1100 | 1024 | 1655 | 395 | 581 | 1076 | 1016 | 394 | 410 | 811 |
| 65-74 | 584 | 686 | 625 | 926 | 309 | 464 | 625 | 637 | 320 | 305 | 447 |
| Seroprevalence (unweighted) |  |  |  |  |  |  |  |  |  |  |  |
| 18-49 | 0.02 | 0.05 | 0.07 | 0.06 | 0.06 | 0.06 | 0.05 | 0.07 | 0.18 | 0.19 | 0.19 |
| 50-64 | 0.03 | 0.06 | 0.05 | 0.05 | 0.05 | 0.05 | 0.04 | 0.05 | 0.09 | 0.18 | 0.14 |
| 65-74 | 0.03 | 0.03 | 0.04 | 0.04 | 0.04 | 0.04 | 0.04 | 0.03 | 0.08 | 0.12 | 0.13 |

(B) Numbers per study
| Study | 2020-03-01 | 2020-04-01 | 2020-05-01 | 2020-06-01 | 2020-07-01 | 2020-08-01 | 2020-09-01 | 2020-10-01 | 2020-11-01 | 2020-12-01 | 2021-01-01 |
| --- | --- | --- | --- | --- | --- | --- | --- | --- | --- | --- | --- |
| Number of samples |  |  |  |  |  |  |  |  |  |  |  |
| Residual Laboratory | 2765 | 2406 | 2356 | 4460 | 0 | 0 | 2320 | 2240 | 0 | 0 | 0 |
| Blood Donors | 584 | 1800 | 1799 | 1800 | 1800 | 2667 | 1797 | 1799 | 1799 | 1808 | 3035 |
| Seroprevalence (unweighted) |  |  |  |  |  |  |  |  |  |  |  |
| Residual Laboratory | 0.03 | 0.05 | 0.06 | 0.05 |  |  | 0.04 | 0.05 |  |  |  |
| Blood Donors | 0.02 | 0.05 | 0.05 | 0.05 | 0.06 | 0.05 | 0.05 | 0.07 | 0.14 | 0.17 | 0.16 |

### 3.2. OBSERVED SEROPREVALENCE AND THE CUMULATIVE PROPORTION INFECTED

On July 5 2020, the cumulative incidence (expressed as a percentage) was estimated at 5.6% (95%CrI 4.9-6.2), 4.0% (95%CrI 3.4-4.7) and 2.4% (95%CrI 1.8-3.2) for the 18-49, 50-64 and 65-74 year-olds, respectively. On 31 January 2021, the cumulative incidence was estimated at 18.9% (95%CrI 17.6- 20.5), 13.5% (95%CrI 11.9-15.4) and 10.7% (95%CrI 8.4-13.1) for the 18-49, 50-64 and 65-74 year-olds, respectively (Fig 1). Overlaid curves of the cumulative incidence by age group are presented in the S3 Fig. The posterior estimates for the Weibull-Bi-exponential distribution representing the sensitivity of the serological tests by age group, clinical severity and time since infection are presented in Fig 2.

**Fig 1:**
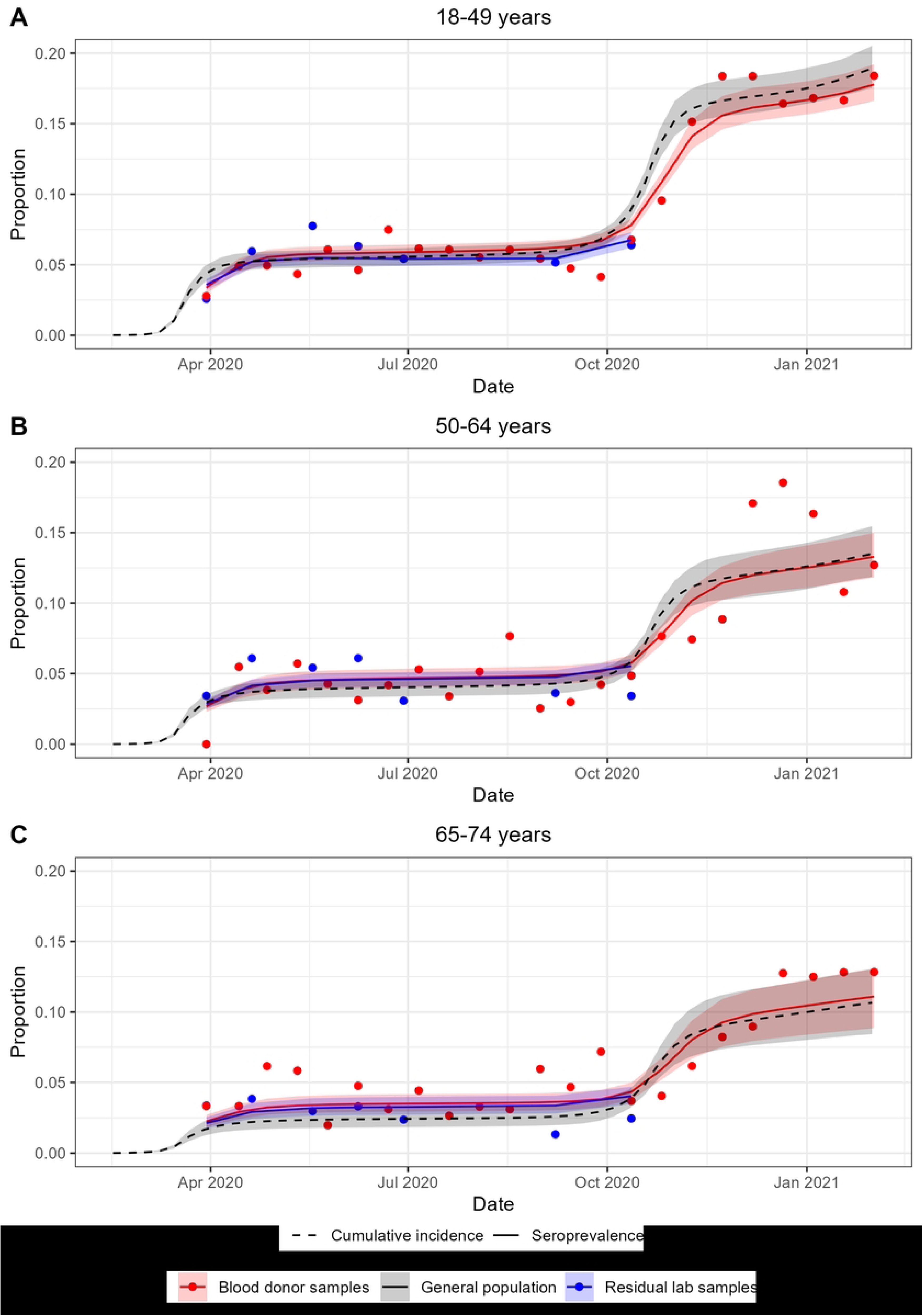
The weighted seroprevalence observed by study by collection round is presented by dots, while the posterior model fit for the seroprevalence and its 95% CrI are presented with lines for both seroprevalence studies. In addition, the cumulative incidence and its 95%CrI are presented for age group 18-49 years (A), 50-64 years (B) and 65-74 years (C).

**Fig 2:**
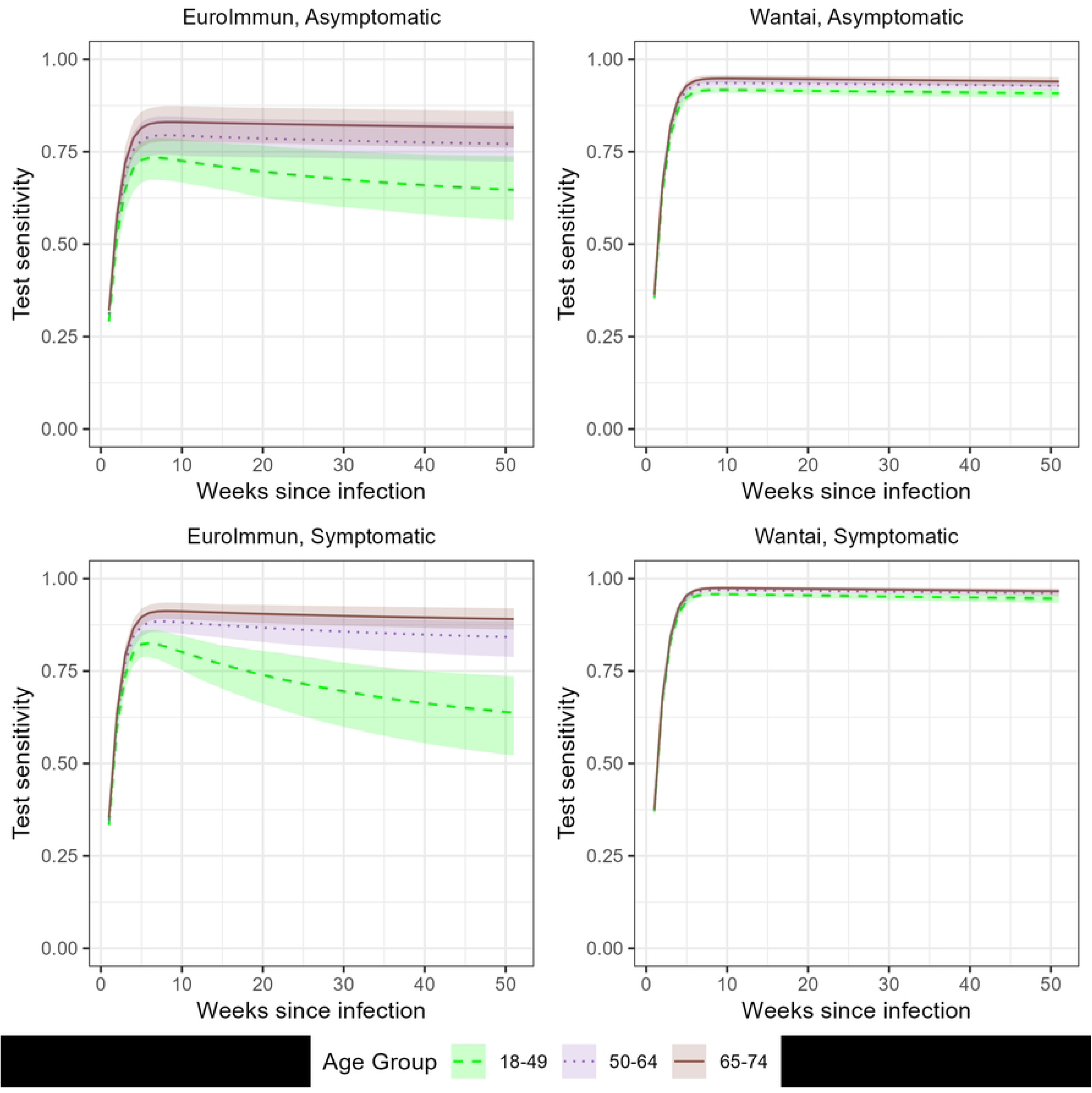
Proportion seropositive after infection. Posterior time-since-infection varying estimates for the test sensitivity of the EuroImmun (left) and Wantai (right) test. For asymptomatic infection (upper) and symptomatic infection (lower) by age group (18-49 years, 50-64 years and 65-74 years).

### 3.3. INFECTIONS, IHR AND IFR

Infections were mainly acquired during the two ‘waves’ (surges of SARS-CoV-2 infections), with few infections in between. While the younger age group was most affected during the first wave, the impact during the second wave was more evenly distributed across all age groups. The proportion infected in the oldest age group however remained smaller than among younger age groups. During the week with the highest incidence of the first wave, starting on March 22, the percentage infected was 2.0% (95% CrI 1.7-2.3), 1.4% (95%CrI 1.2-1.7) and 0.7% (95% CrI 0.5-1.0) in 18-49, 50-64 and 65-74 year-olds, respectively. During the week with the highest incidence of the second wave, starting October 25, the weekly proportion infected was 2.8% (95% CrI 2.3-3.3), 2.1% (95%CrI 1.7-2.7) and 1.6% (95% CrI 1.2-2.1) in 18-49, 50-64 and 65-74 year-olds, respectively (Fig 3).

**Fig 3:**
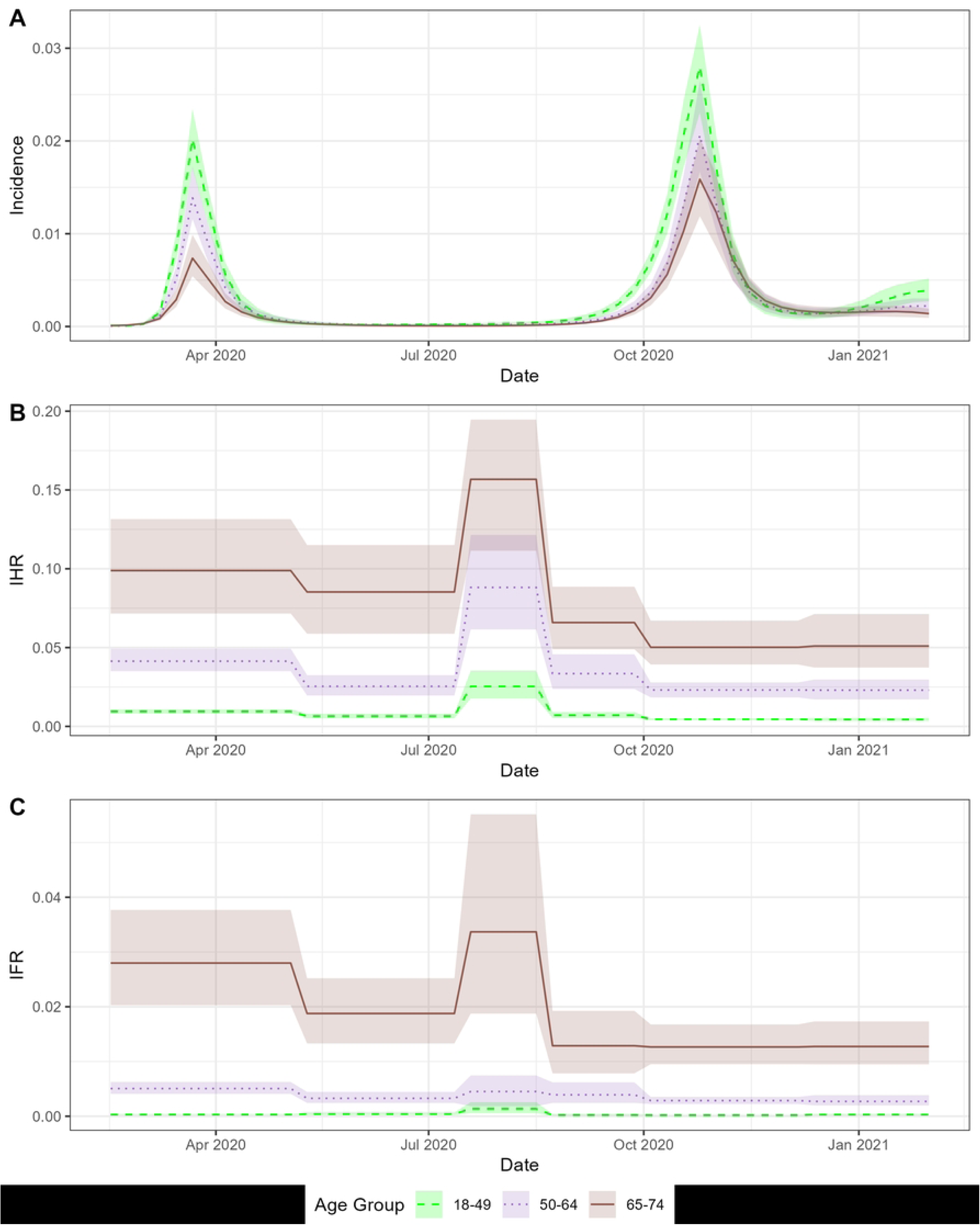
The proportion of the population infected by week (A), the Infection Hospitalization Rate (IHR, B) and the Infection Fatality Rate (IFR, C).

The IHR declined over the study period, during the first lockdown the percentage hospitalized after infection was estimated at 9.9% (95%CrI 7.2-13.2), while it was estimated at 5.1% (95%CrI 3.8-7.1) in the period following the second lockdown in the oldest age group. For persons 50 to 64 years old the IHR went from 4.1% (95%CrI 3.5-4.9) to 2.3% (95%CrI 1.7-3.0) and in the youngest age group, the percentage declined from 0.9% (95%CrI 0.8-1.0) to 0.4% (95%CrI 0.3-0.5). A similar trend in which the rates were significantly reduced over 2020 was observed for the IFR. For persons 65 to 74 years old, the IFR decreased from 2.8% (95%CrI 2.0-3.8) to 1.3% (95%CrI 0.9-1.7), for persons 50 to 64 years old, it decreased from 0.5% (95%CrI 0.4-0.6) to 0.3% (95%CrI 0.2 to 0.4). For persons 18 to 49 years, the IFR remained at a very low percentage: 0.03% (95%CrI 0.02-0.04) to 0.03% (95%CrI 0.02-0.04). During the period from mid-July to end of August, both the IHR and IFR increased.

### 3.4. MODEL GOODNESS OF FIT

The model provided acceptable posterior predictive intervals for the fitted data: seroprevalence results of RLS and BDS, the age-specific hospitalizations and deaths (S4 Fig). Additional variation was observed for the hospitalization time series with 11 observations outside of the 95% prediction intervals out of 153 (7.2%). The posterior estimate for the overdispersion parameter was smallest (most overdispersion) for the 18-49 year-olds of the RLS (3638, 95%CrI 1202-8022).

Graphical overviews by age group of the time series of infections, cases, intakes and deaths are provided in S5 Fig. Direct case counts were not proportional to the estimated number of infections over 2020. In the first half of 2020, testing capacity was too limited for case counts to represent the magnitude of the surge relative to later case ascertainment. In the second half of 2020 a change in testing strategy; amongst others the testing of asymptomatic high-risk contacts was no longer mandated, accelerated the peak in case counts compared to the peak in infections for the youngest age group.

The discrete delay distribution for hospitalizations peaked in the first week. Of those infections that result in hospitalization, 67% (95%CrI 59%-76%, 18-49y), 76% (95%CrI 64%-86%, 50-64y) and 89% (95% 78%-97%) did so within the week of infection. In comparison, only few infections resulted in death within the same week. For the youngest age group, deaths mostly occurred two weeks after infection. While for the older age groups deaths mostly occurred in the week after infection (S1 Table).

Our sensitivity analysis (SA) to assess robustness to the distribution and informative priors used for the tests’ sensitivity (S6 Fig) resulted in the following two main observations. The SA-sensitivity was higher for younger age groups resulting in a decrease in estimated incidence in the SA compared to the main analysis (17.4% compared to 19%). Second, the fit to the seroprevalence data was worse in the SA, including a higher estimate for the overdispersion parameter, compared to the main analysis.

## 4. Discussion

Our work contributes to an ongoing effort to estimate the incidence of SARS-CoV-2 infections during the initial years of the pandemic. We combined data from two different repeated cross-sectional seroprevalence studies with surveillance data on COVID-19 hospitalizations and deaths. In addition, we included informative priors for the specificity and time-since-infection varying sensitivity of the serological tests. Posterior incidence estimates showed two waves in 2020, with little transmission in between. We estimated that 16.2% of the general Belgian population aged 18 to 74 years old had been infected by the start of 2021. The first wave mainly affected those under 50 years old. The second wave affected age groups more equally and was associated with lower age-specific infection fatality and hospitalization rates.

### 4.1. SEROPREVALENCE RESULTS

With seroprevalence estimates around 16% for the general adult population, Belgium ranked high among the list of countries affected by COVID-19 in 2020. A systematic review of sero-epidemiological studies estimated the worldwide seroprevalence at 4.5% by December 2020 [10]. Among European countries, Germany, Norway, Denmark and Iceland can be classified as having low seroprevalence with 1.7% (November [32]), 0.9% (December [40]), 2.2% (October [41]) and 0.9% (June [42]), respectively. European countries reporting higher seroprevalence were Spain, the Netherland and England with 9.8% (November 2020 [43]), 12.2% (February 2021 [44]) and 12.1% (December 2020 [45]), respectively. A direct comparison however is not possible given the limitations of seroprevalence studies presented earlier.

While we did report age group-specific estimates, we did not explore incidences within specific settings. Other Belgian seroprevalence studies have explored specific settings and populations and explored factors correlated to seropositivity. Slightly higher seroprevalence estimates following the same overall trends have been reported for Belgian healthcare workers. After the first wave seroprevalence was estimated around 7% in healthcare workers [12,46]. Studies in healthcare workers reported specific workplace (e.g. working in a general COVID-19 ward, but not intensive care), personal protective equipment and non-occupational related risk factors (e.g. household exposure, high population incidence in the area, …) as associated with seropositivity [12,47–49]. These observations led researchers to conclude that preventive measures were effective in reducing occupational risk. The observed seroprevalence in nursing home residents and staff, estimated at 17-21% shortly after the first wave [50], however illustrates high transmission in this specific healthcare setting. Seroprevalence trends within and between nursing homes are hard to interpret given the high mortality in seropositive residents [51]. A higher fraction of dependent residents [52] and residents with dementia [53] have been associated with SARS-CoV-2 circulation. Finally, seroprevalence estimates in children 6 years and older were lower or comparable to the general population [54–56].

### 4.2. IFR/IHR

Our IHR and IFR estimates and their evolution over time are in accordance with results from meta-analyses which likewise reported a decrease over 2020 [16,57,58]. These findings have been linked to improvements in treatment [59]. Discussion remains about the exact trend over 2020 [60]. Because of temporal changes in hospital occupancy and admission protocols [61], short-term trends might deviate from the long-term trend. We observed an increase in IHR and IFR in August 2020 coinciding with a prolonged heatwave. This heatwave was associated with considerable excess mortality [62]. A heatwave can directly affect IHR and IFR by increasing patient vulnerability. Alternatively, given the relatively low infection incidence during this period, even minor misclassification in COVID-19- associated hospitalizations or deaths could disproportionately affect IHR and IFR calculations.

Molenberghs et al. [63] estimated lower IFR for the first wave (March-May 2020) including the same mortality data as this study, the difference therefore is in the estimated number of infections. Molenberghs et al. obtained incidences from a stochastic compartmental model [64]. This model, calibrated to, amongst others, the RLS-study, estimated cumulative incidences at 6.9% on May 4, 2020. The higher estimates for the cumulative incidence might result from the assumption of perfect specificity by the compartmental model. The assumed lack of false positives partly explains why the previously published IFR-estimates are below this study’s IFR estimates. Imperfect test specificity can substantially inflate observed seroprevalence, particularly when actual infection rates are low. While the sensitivity of the tests used was generally lower than their specificity, this doesn’t inherently mean observed seroprevalence underestimates cumulative incidence. The relationship hinges on the true prevalence level, which dictates whether false positives (due to imperfect specificity) or false negatives (due to imperfect sensitivity) have a greater numerical impact. In lower-prevalence groups false positives will inflate the observed rate. However, in our highest-incidence group (18-49 years) later in 2020, the prevalence became high enough for the number of false negatives to exceed false positives.

### 4.3. METHODS

Given the test-specific and time-varying sensitivity associated with qualitative immuno-assays, further complicated by the interaction with characteristics of the cohort and the infections within the cohort, considerable data synthesis is necessary to combine seroprevalence data collected through different studies from different cohorts to estimate incidence. Several approaches have been suggested. This study is different from studies such as those using REMEDID [65] and the works by IHME [16] and Shioda et al. [66]. In our study, seroprevalence data is directly included. Other studies have taken a stepwise, indirect approach: first estimating IHR/IFR from seroprevalence data and then estimating incidence from IHR/IFR and hospitalizations and deaths. The first step, estimation of IFR, might include temporal variation, but often temporal homogeneity is assumed [67]. We decided to include all available data directly within the same unified fully Bayesian statistical framework. A second difference is the inclusion of age and severity-specific sensitivity estimates. Especially severity of infection is a major factor affecting the sensitivity of serological testing [68]. We included sensitivity using a Weibull-Bi-exponential distribution with informative priors for the distributional parameters. Shioda et al. [66] proposed a Weibull-Weibull distribution. Other have used single Weibull distributions [66,69,70] and constant exponential rates [71–73] for modelling these immunity kinetics. In addition to the distributions themselves, the estimation of distributional parameters is a topic of discussion. Shioda et al. estimated these parameters from the seroprevalence data itself. Similar to Takahashi et al. [67], Kadelka et al. [14] and others [74,75], we considered it essential to estimate these parameters, representing individual-level immune dynamics, from longitudinal data on serological testing. This difference in approach has been evaluated by Kadelka et al. [14]. They revised a cumulative incidence estimate by Buss et al. [71] of 76% to 47.6%. The former estimated the seroreversion rate directly from the serosurvey data, while the latter included longitudinal antibody data. Finally, our approach allowed for the delay distributions to be estimated from the data. The delay distribution did not have to be defined a priori, which was a requirement in the framework of Shioda et al. [66] and in REMEDID [65].

The study by Barber et al. [16] deserves some further attention as it largely shares both the objective and data included in this study. While there were differences: seroprevalence was included indirectly, specificity of serological tests was ignored and the included decrease in sensitivity of the EuroImmun test over time was much more aggressive, the results reported for Belgium are comparable to ours. They estimated around 2 million infections at the start of 2021 (∼18% of the population infected in 2020), accumulated over two waves in 2020 of which the second was the larger wave. Their IFR estimate and trend was also comparable to ours.

### 4.4. LIMITATIONS

We observed overdispersion associated with the seroprevalence studies. It is important to note that these samples remain samples of convenience. Little info is available on the persons present in these samples. We divided them into wide age groups without further differentiation, leaving multiple points on which they might differ from the general population or be heterogeneous over time. This is well illustrated by Herzog et al. [17] who discussed the possible contribution of self-confinement of the typical healthcare seeking person on participation to the earlier collection rounds of the RLS compared to later rounds. Whatever the mechanisms underlying the observed overdispersion, they are not well understood and require further investigation to improve inference from seroprevalence studies.

Our use of a beta-binomial model for seroprevalence allowed us to assess variability beyond simple sampling effects (overdispersion). While our model priors were set to favor minimal overdispersion initially, they were flexible enough to adapt to the data. The finding of significant overdispersion in the results confirms that the observed seroprevalence varied more than expected based solely on modelled infection rates and sample sizes. This excess variability likely arises from limitations inherent in the seroprevalence studies. These include potential unmeasured cohort heterogeneity, spatial effects, differing study protocols (BDS vs. RLS), and selection biases associated with convenience sampling, none of which could be fully explored with the available data. Effectively controlling for such sources of variability and bias remains a critical aspect of robust seroprevalence study design [76].

This study uses a spline to model the underlying incidence of infections. This incidence is constrained by the spline’s structure. We explored different possible splines by fitting them to advanced mortality and hospitalization time series. Other possibilities have not been explored in this paper. Case data is presented alongside the model outputs (S5 Fig), but it was not used in the model fit or for deciding on possible knot placement. Insufficient testing capacity in the first half of 2020 and changes in testing strategy when incidence increased and exceeded testing capacity again during the fall of 2020 created temporal changes in the case ascertainment that are not investigated in this paper. In addition, the analysis might be sensitive to the periods used for IHR and IFR.

PCR-confirmed infection is used in this paper as a proxy for SARS-CoV-2 infection. Much of the work on test sensitivity and specificity, but also work on IFR and IHR is based on PCR-confirmed infections. Translating modelled infection timing to potential PCR-confirmation timing would require an additional step incorporating delays, test sensitivity, and specificity associated with PCR testing itself.

By estimating that 16.2% of Belgian adults were infected with SARS-CoV-2 in 2020 – mainly during two distinct waves with low transmission between them – our findings offer significant public health insights. Understanding the true scale of infection, the age-related risks, and the decline in severity over the year is vital for retrospectively evaluating the effectiveness of public health measures implemented in Belgium and for comprehending the population immunity landscape preceding vaccination. Furthermore, this research highlights the importance for public health policy of using integrated data approaches. Accurately estimating infection incidence, burden, and delays, while accounting for data limitations like test performance, is essential for effective resource allocation, strategic planning, and enhanced preparedness for future public health emergencies.

## Data Availability

Data are available on Zenodo (http://doi.org/10.5281/zenodo.4664403) Belgian COVID-19 data can be requested through: https://catalog.hda.belgium.be/search?page=1&query=covid&unionType=0

https://catalog.hda.belgium.be/search?page=1&query=covid&unionType=0

## ACKNOWLEDGMENTS

We acknowledge Caroline Rodeghiero, Fabienne Jurion, Christophe Van den Poel, Sara Vande Kerckhove, Elfriede Heerwegh, Elien De Vits, Jessie Claessens, Veerle Van Melle and Martine Clinet for their excellent work in managing and analysing the thousands of plasma samples. We also thank An Muylaert and Dominique Goossens for coordinating the study at the side of the red cross (BDS study).

We acknowledge the Belgian laboratories that voluntarily collected sera and data for this study: Algemeen Medisch Laboratorium (AML, Antwerpen), Laboratoire Luc OLIVIER (Fernelmont), Declerck Klinisch Laboratorium (Ardooie), Klinisch Labo RIGO (Genk), Labo Anacura/Nuytinck (Evergem), Labo Somedi (Heist-op-den-Berg), Labo LBS (Brussels), Laboratoire Bauduin (Enghien), Medisch labo Bruyland (Kortrijk), Synlab (Luik).

---

**S1 Fig.**
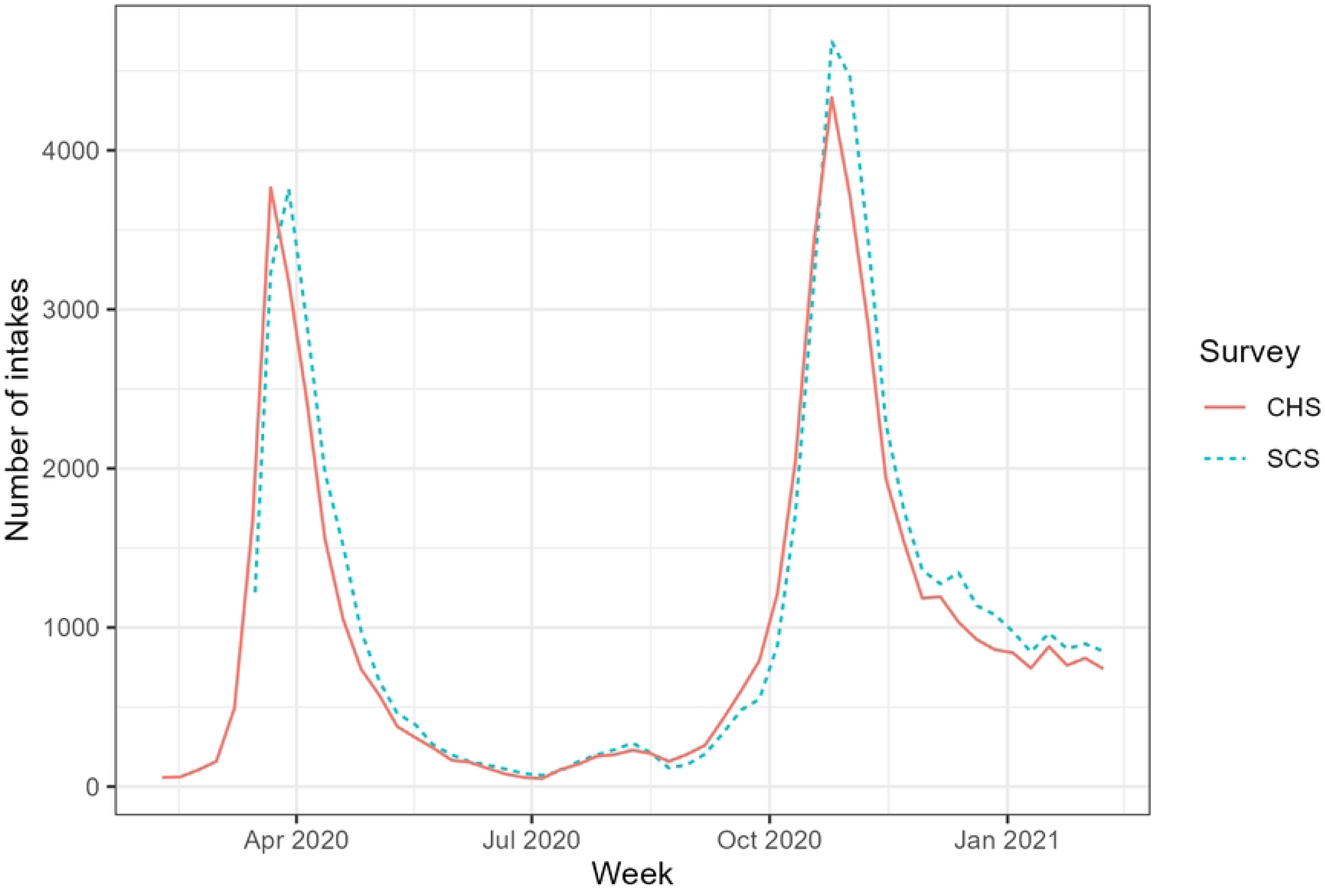
: The number of intakes reported through the Surge Capacity Survey (SCS) and the Clinical Hospital Survey (CHS). The SCS and the CHS are two distinct surveys. Participating in the SCS was mandatory and the number of new intakes had to be reported within 24 hours. In addition to surveillance purposes, the survey helped manage capacity and transfers. No individual data were collected. The CHS was a voluntary survey that collected data on the patient profile, length of stay, clinical characteristics at presentation and during the hospital stay and outcomes. While voluntary, the participation to this survey was very good throughout 2020.

**S2 Fig.**
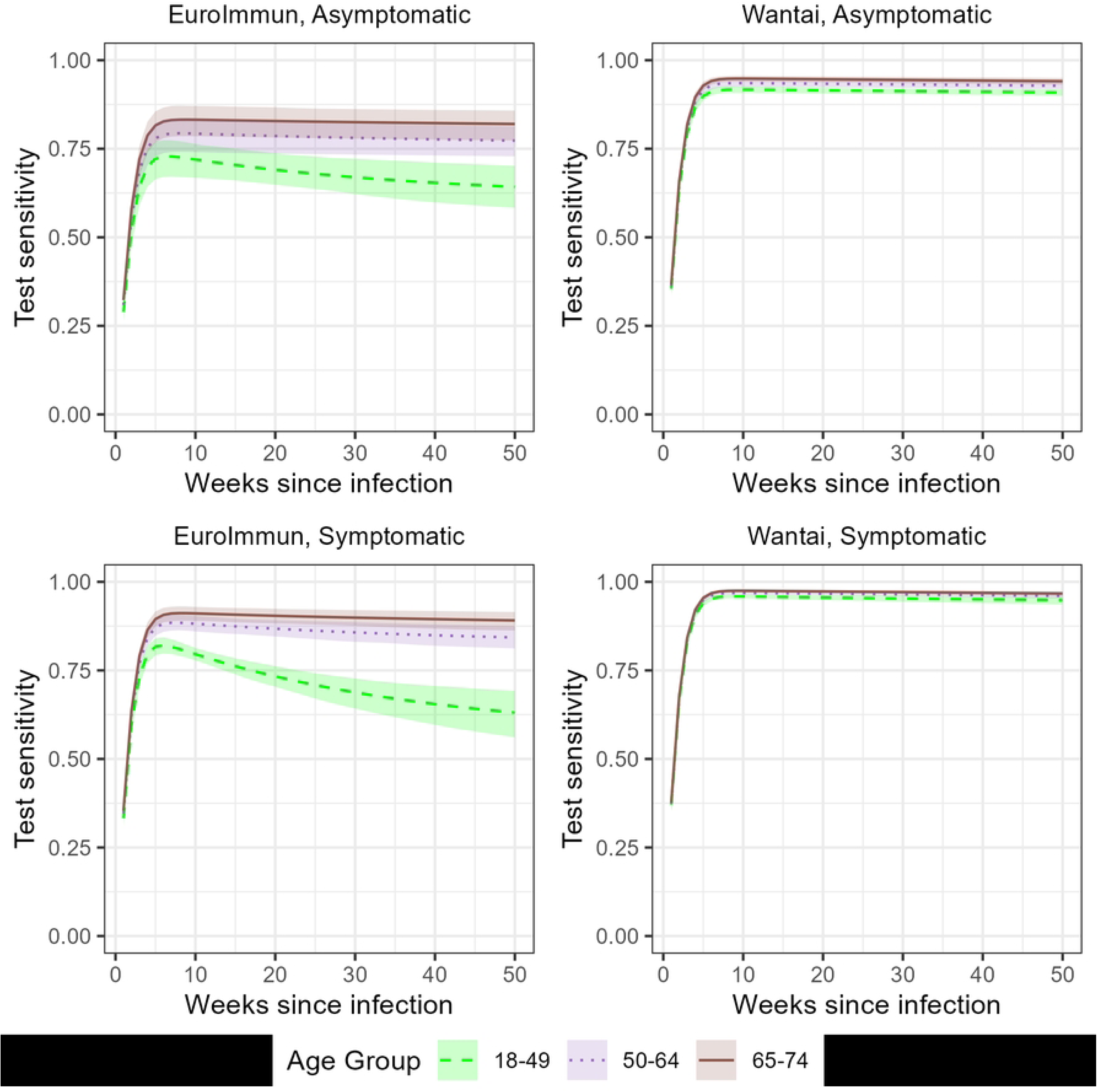
: The scaled Weibull-Weibull distribution resulting from the informative priors for the EuroImmun test (left) and the Wantai test (right) for asymptomatic infection (upper) and symptomatic infections (lower). In previous work, we combined data on seroconversion and reversion from literature and Belgium’s laboratory SARS-CoV-2 testing to obtain parametric curves for the scaled seroconversion and seroreversion.

**S3 Fig.**
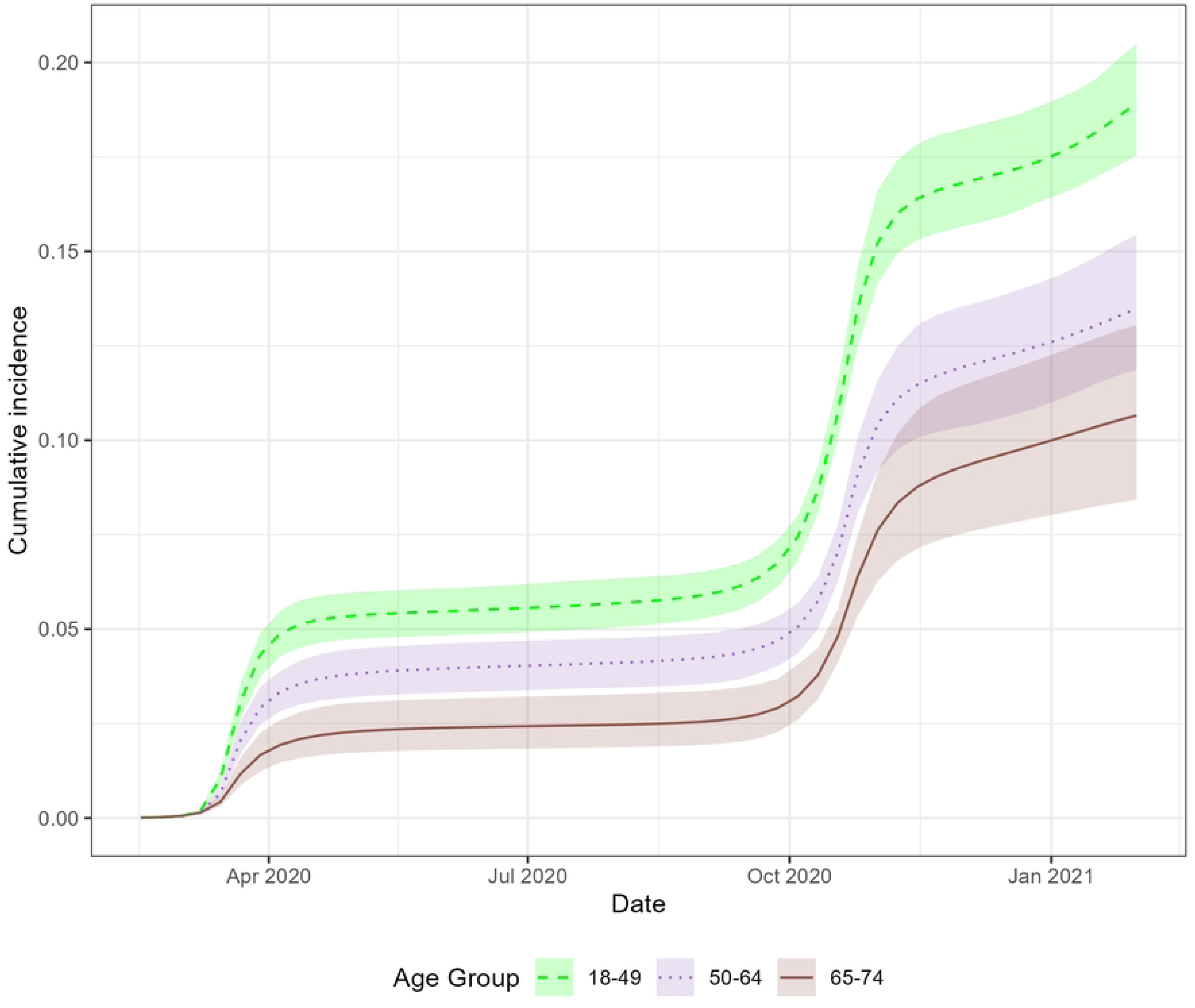
: The cumulative proportion infected over time by age group (18 to 49 years, 50 to 64 years and 65 to 74 years of age).

**S4 Fig.**
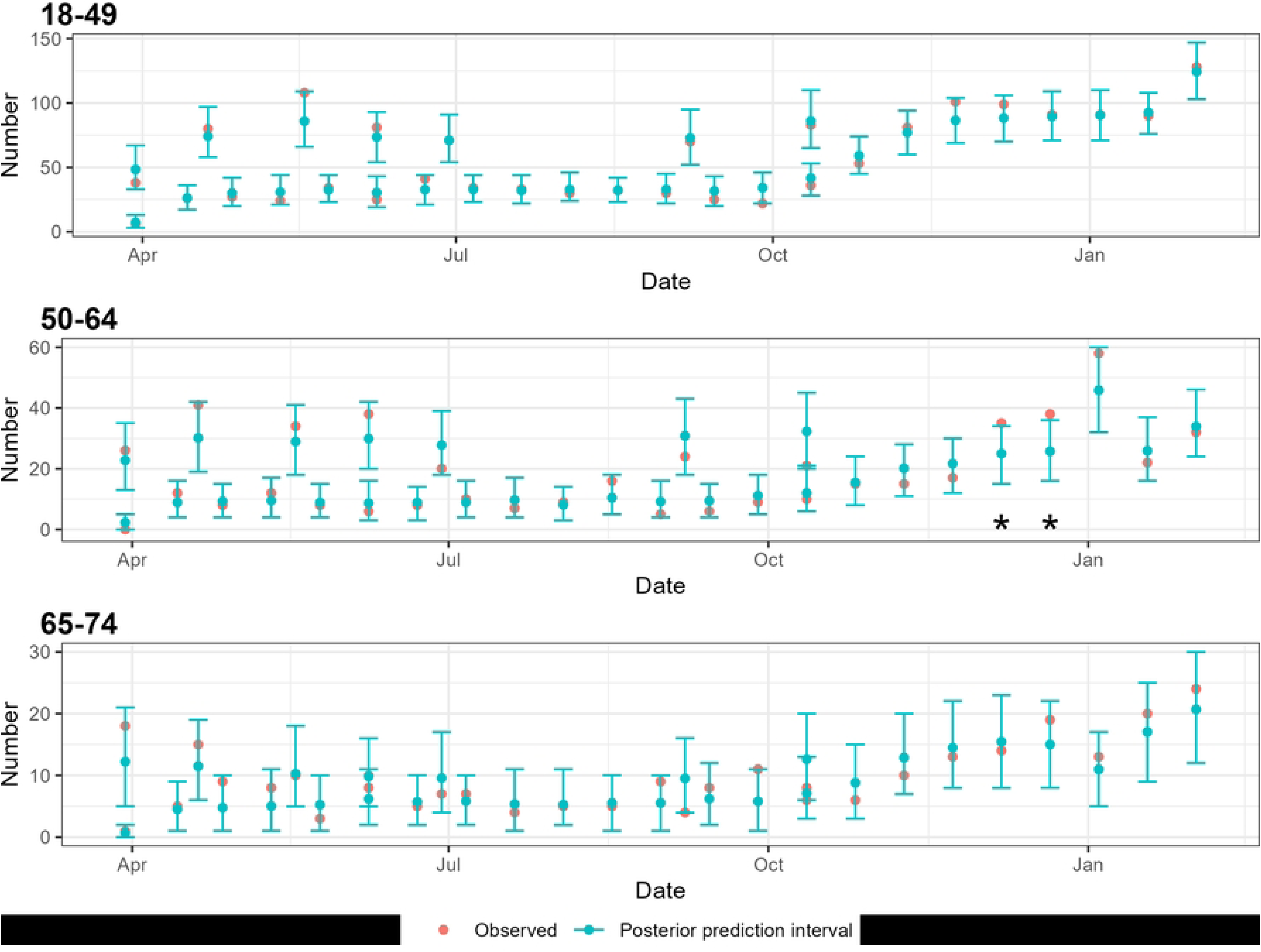
(A): Posterior 95% Predictive Intervals from the beta-binomial distribution for the seroprevalence by age group (18 to 49 years, 50 to 64 years and 65 to 74 years of age). Both the BDS and the RLS seroprevalence study are presented. Stars mark those observations outside of the 95% predictive intervals.

**S4 Fig.**
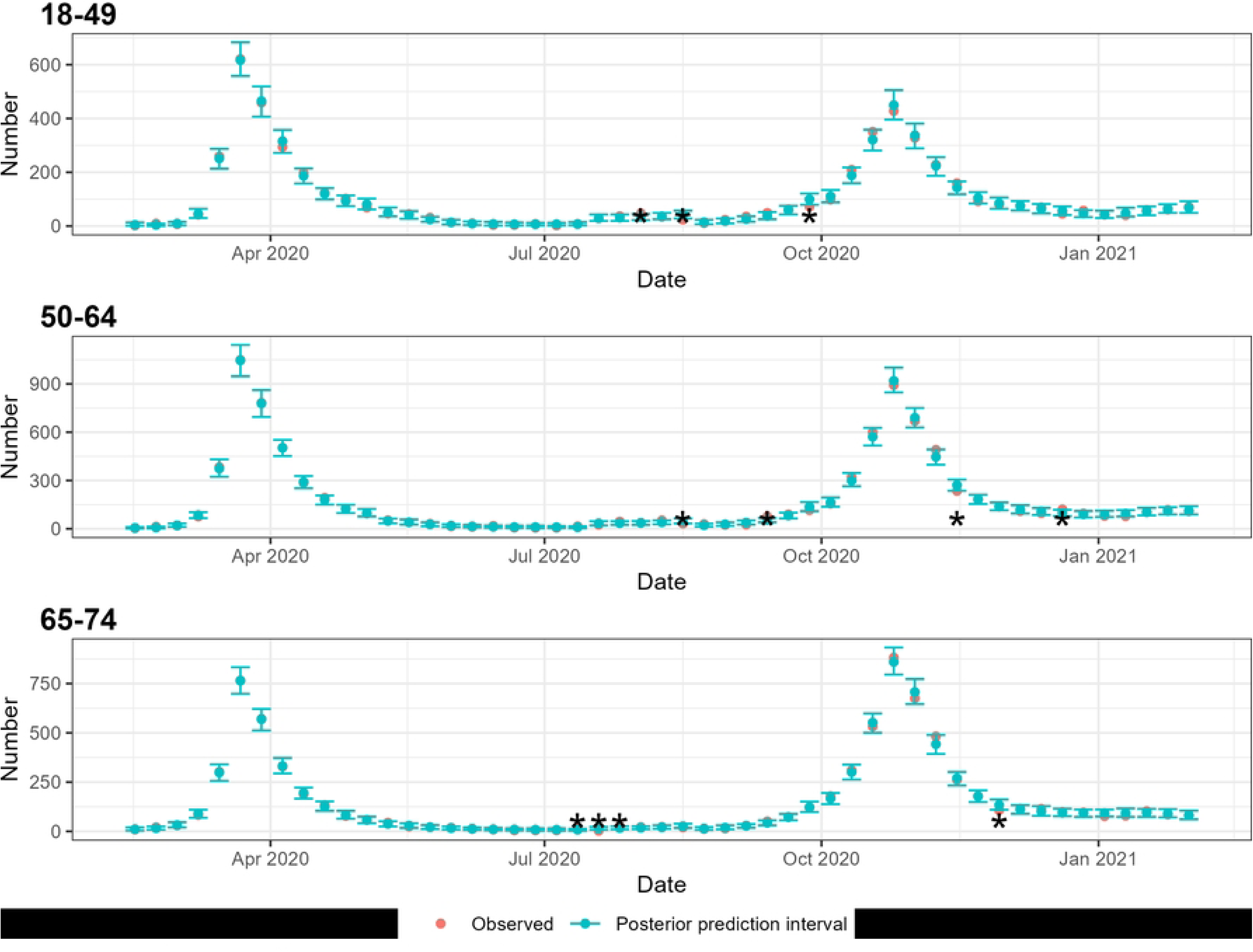
(B): Posterior 95% Predictive Intervals from the binomial distribution for the weekly number of hospitalizations in the CHS by age group (18 to 49 years, 50 to 64 years and 65 to 74 years of age). Stars mark those observations outside of the 95% predictive intervals.

**S4 Fig.**
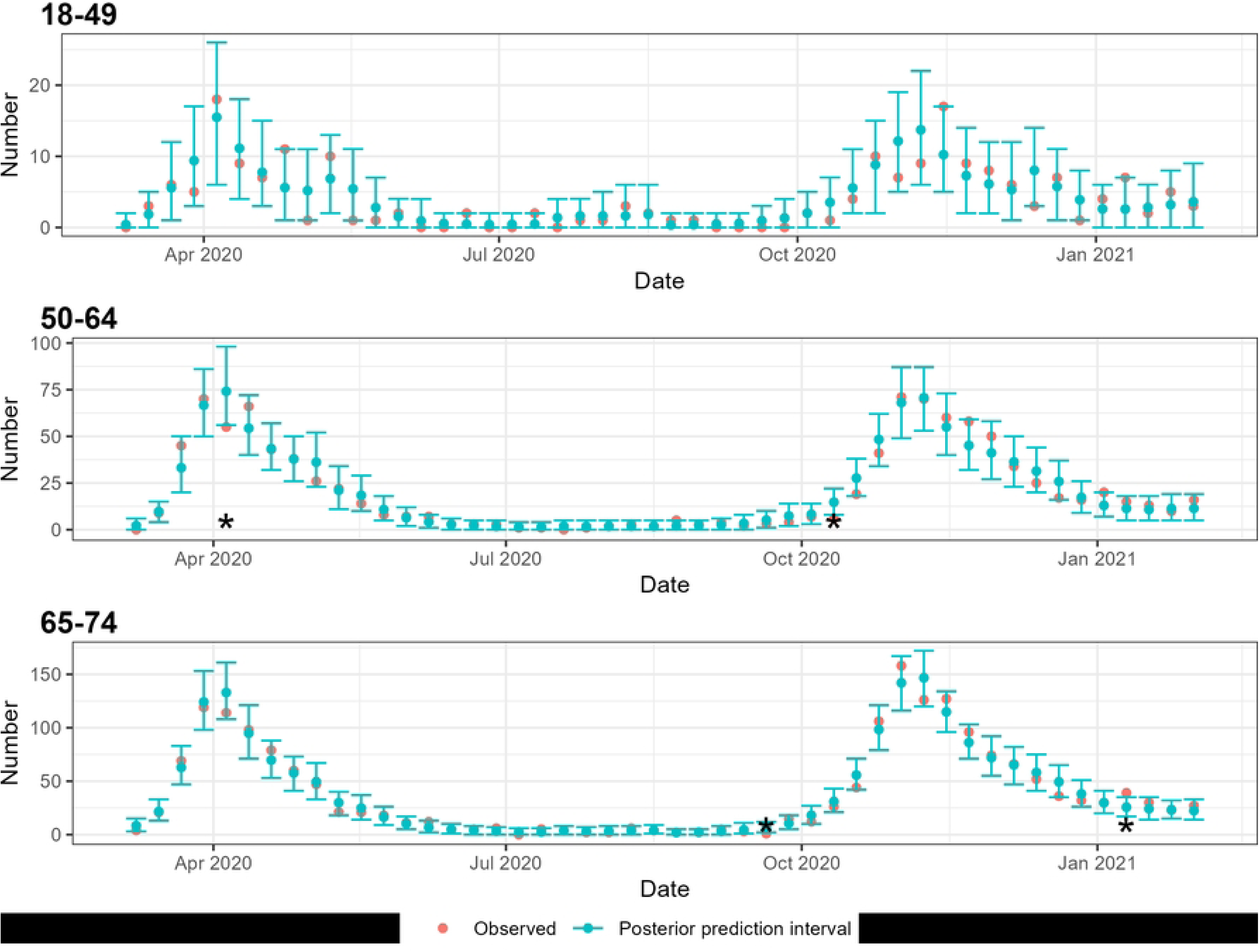
(C): Posterior 95% Predictive Intervals from the binomial distribution for the weekly number of deaths by age group (18 to 49 years, 50 to 64 years and 65 to 74 years of age). Stars mark those observations outside of the 95% predictive intervals.

**S5 Fig.**
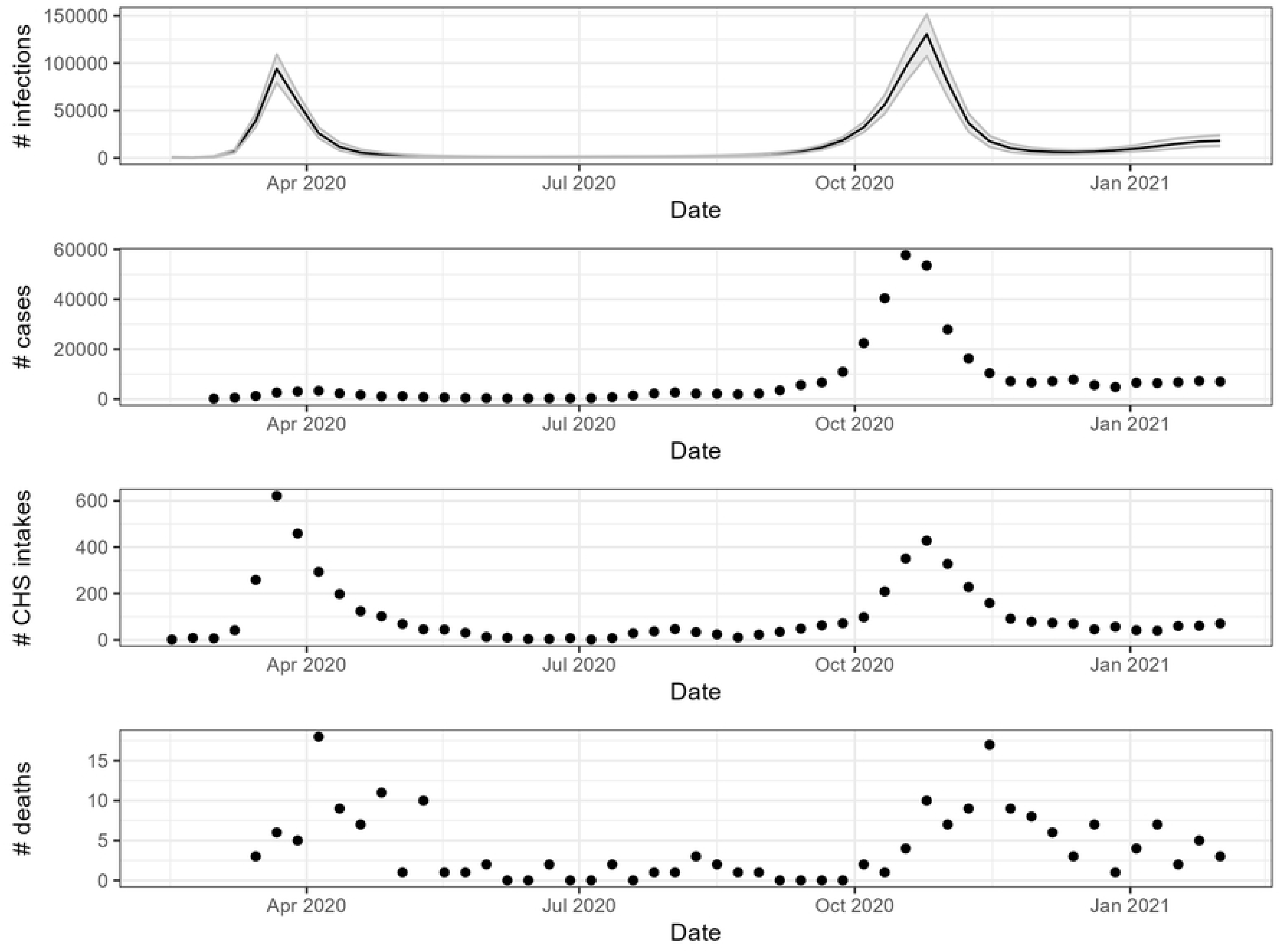
(A): Time series for the weekly number of infections (including 95% CI), cases, Clinical Hospital Survey (CHS) intakes and deaths in the age group 18 to 49 years.

**S5 Fig.**
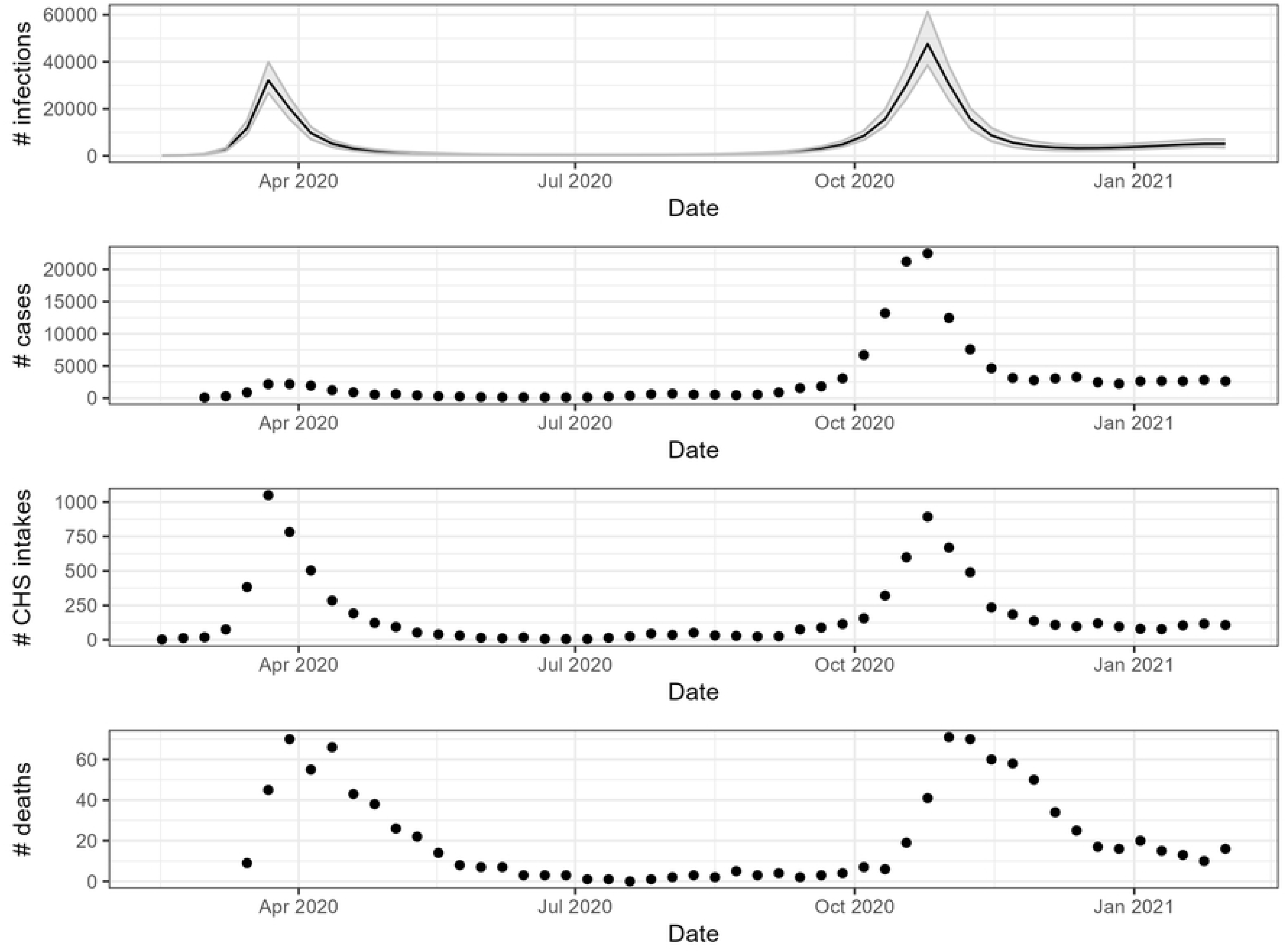
(B): Time series for the weekly number of infections (including 95% CI), cases, Clinical Hospital Survey (CHS) intakes and deaths in the age group 50 to 64 years.

**S5 Fig.**
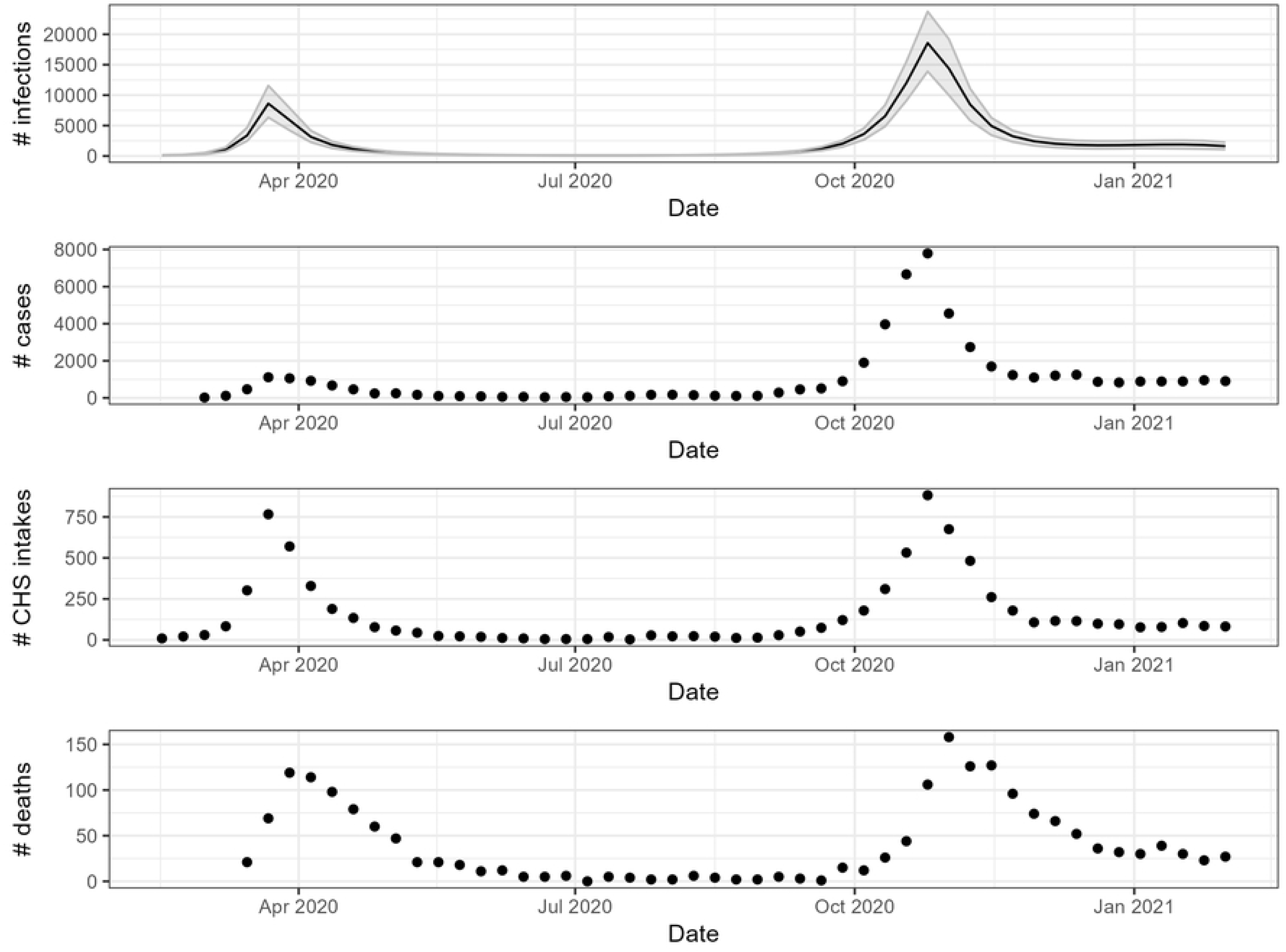
(C): Time series for the weekly number of infections (including 95% CI), cases, Clinical Hospital Survey (CHS) intakes and deaths in the age group 65 to 74 years.

**S6 Fig.**
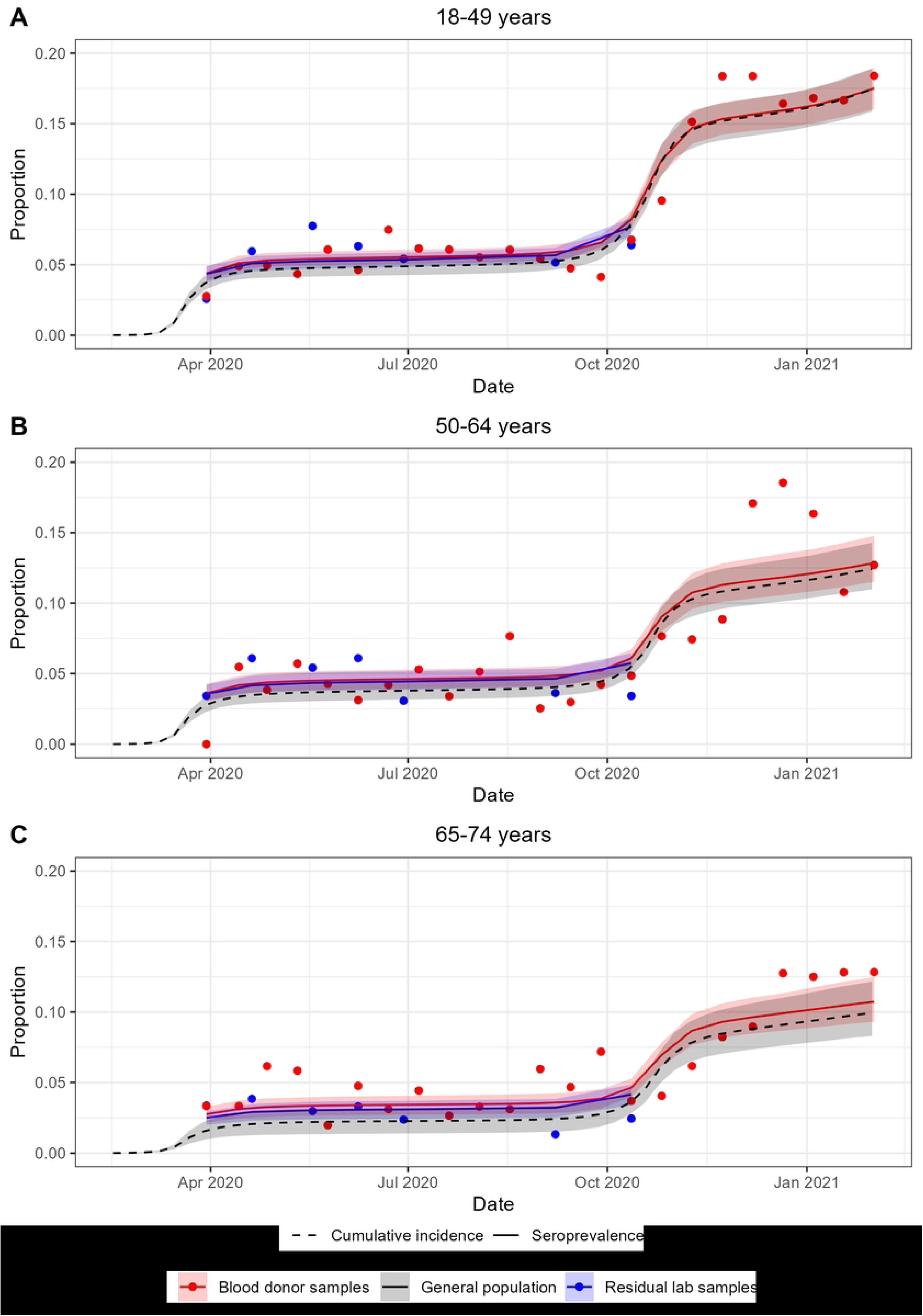
: Sensitivity analysis to assess the impact of the distributions for the serological tests’ sensitivity. In the sensitivity analysis, the sensitivity of the serological tests has one, age- and time-invariant, sensitivity estimate per test.

## REFERENCES

1. Sierra NB, Bossuyt N, Braeye T, Leroy M, Moyersoen I, Peeters I, et al. All-cause mortality supports the COVID-19 mortality in Belgium and comparison with major fatal events of the last century. Arch Public Health. 2020;78(1):1–8.

2. Sciensano. Epistat - Belgian COVID-19 opendata [Internet]. [cited 2020 Sep 12]. Available from: https://epistat.wiv-isp.be/covid/

3. Meurisse M, Lajot A, Dupont Y, Lesenfants M, Klamer S, Rebolledo J, et al. One year of laboratory- based COVID-19 surveillance system in Belgium: main indicators and performance of the laboratories (March 2020-21). Arch Public Health Arch Belg Sante Publique. 2021 Oct 7;79(1):188.

4. Duysburgh E, Mortgat L, Barbezange C, Dierick K, Fischer N, Heyndrickx L, et al. Persistence of IgG response to SARS-CoV-2. Lancet Infect Dis. 2021 Feb;21(2):163–4.

5. To KKW, Tsang OTY, Leung WS, Tam AR, Wu TC, Lung DC, et al. Temporal profiles of viral load in posterior oropharyngeal saliva samples and serum antibody responses during infection by SARS-CoV-2: an observational cohort study. Lancet Infect Dis. 2020 May;20(5):565–74.

6. Xu X, Sun J, Nie S, Li H, Kong Y, Liang M, et al. Seroprevalence of immunoglobulin M and G antibodies against SARS-CoV-2 in China. Nat Med. 2020 Aug;26(8):1193–5.

7. Wajnberg A, Amanat F, Firpo A, Altman DR, Bailey MJ, Mansour M, et al. Robust neutralizing antibodies to SARS-CoV-2 infection persist for months. Science. 2020 Dec 4;370(6521):1227–30.

8. Rostami A, Sepidarkish M, Leeflang MMG, Riahi SM, Nourollahpour Shiadeh M, Esfandyari S, et al. SARS-CoV-2 seroprevalence worldwide: a systematic review and meta-analysis. Clin Microbiol Infect Off Publ Eur Soc Clin Microbiol Infect Dis. 2021 Mar;27(3):331–40.

9. Hanson KE, Caliendo AM, Arias CA, Englund JA, Hayden MK, Lee MJ, et al. Infectious Diseases Society of America Guidelines on the Diagnosis of COVID-19:Serologic Testing. Clin Infect Dis Off Publ Infect Dis Soc Am. 2020 Sep 12;

10. Bergeri I, Whelan MG, Ware H, Subissi L, Nardone A, Lewis HC, et al. Global SARS-CoV-2 seroprevalence from January 2020 to April 2022: A systematic review and meta-analysis of standardized population-based studies. PLOS Med. 2022 Nov 10;19(11):e1004107.

11. Lohse S, Sternjakob-Marthaler A, Lagemann P, Schöpe J, Rissland J, Seiwert N, et al. German federal-state-wide seroprevalence study of 1st SARS-CoV-2 pandemic wave shows importance of long-term antibody test performance. Commun Med. 2022 May 18;2(1):1–12.

12. Mortgat L, Verdonck K, Hutse V, Thomas I, Barbezange C, Heyndrickx L, et al. Prevalence and incidence of anti-SARS-CoV-2 antibodies among healthcare workers in Belgian hospitals before vaccination: a prospective cohort study. BMJ Open. 2021 Jun 1;11(6):e050824.

13. ROGAN WJ, GLADEN B. ESTIMATING PREVALENCE FROM THE RESULTS OF A SCREENING TEST. Am J Epidemiol. 1978 Jan 1;107(1):71–6.

14. Kadelka S, Bouman JA, Ashcroft P, Regoes RR. Correcting for Antibody Waning in Cumulative Incidence Estimation From Sequential Serosurveys. Am J Epidemiol. 2024 May 7;193(5):777–86.

15. Vaughan A, Duffell E, Freidl GS, Lemos DS, Nardone A, Valenciano M, et al. Systematic review of seroprevalence of SARS-CoV-2 antibodies and appraisal of evidence, prior to the widespread introduction of vaccine programmes in the WHO European Region, January–December 2020. BMJ Open. 2023 Nov 1;13(11):e064240.

16. Barber RM, Sorensen RJD, Pigott DM, Bisignano C, Carter A, Amlag JO, et al. Estimating global, regional, and national daily and cumulative infections with SARS-CoV-2 through Nov 14, 2021: a statistical analysis. The Lancet. 2022 Jun 25;399(10344):2351–80.

17. Herzog SA, Bie JD, Abrams S, Wouters I, Ekinci E, Patteet L, et al. Seroprevalence of IgG antibodies against SARS-CoV-2 – a serial prospective cross-sectional nationwide study of residual samples, Belgium, March to October 2020. Eurosurveillance. 2022 Mar 3;27(9):2100419.

18. Aantal bloeddonoren en gezondheidswerkers met antistoffen tegen coronavirus stijgt [Internet]. sciensano.be. [cited 2025 Mar 21]. Available from: https://www.sciensano.be/nl/pershoek/aantal-bloeddonoren-en-gezondheidswerkers-met-antistoffen-tegen-coronavirus-stijgt

19. Gededzha MP, Mampeule N, Jugwanth S, Zwane N, David A, Burgers WA, et al. Performance of the EUROIMMUN Anti-SARS-CoV-2 ELISA Assay for detection of IgA and IgG antibodies in South Africa. PLOS ONE. 2021 Jun 23;16(6):e0252317.

20. Bailie CR, Tseng YY, Carolan L, Kirk MD, Nicholson S, Fox A, et al. Trend in Sensitivity of Severe Acute Respiratory Syndrome Coronavirus 2 (SARS-CoV-2) Serology One Year After Mild and Asymptomatic Coronavirus Disease 2019 (COVID-19): Unpacking Potential Bias in Seroprevalence Studies. Clin Infect Dis Off Publ Infect Dis Soc Am. 2022 Jan 13;75(1):e357–60.

21. Perez-Saez J, Zaballa ME, Yerly S, Andrey DO, Meyer B, Eckerle I, et al. Persistence of anti- SARS-CoV-2 antibodies: immunoassay heterogeneity and implications for serosurveillance. Clin Microbiol Infect. 2021 Nov 1;27(11):1695.e7-1695.e12.

22. Dehgani-Mobaraki P, Zaidi AK, Yadav N, Floridi A, Floridi E. Longitudinal observation of antibody responses for 14 months after SARS-CoV-2 infection. Clin Immunol. 2021 Sep 1;230:108814.

23. Gallais F, Gantner P, Bruel T, Velay A, Planas D, Wendling MJ, et al. Evolution of antibody responses up to 13 months after SARS-CoV-2 infection and risk of reinfection. EBioMedicine. 2021 Sep;71:103561.

24. Petersen MS, Pérez-Alós L, Armenteros JJA, Hansen CB, Fjallsbak JP, Larsen S, et al. Factors influencing the immune response over 15 months after SARS-CoV-2 infection: A longitudinal population-wide study in the Faroe Islands. J Intern Med. 2023;293(1):63–81.

25. Hønge BL, Hindhede L, Kaspersen KA, Harritshøj LH, Mikkelsen S, Holm DK, et al. Long-term detection of SARS-CoV-2 antibodies after infection and risk of re-infection. Int J Infect Dis. 2022 Mar 1;116:289–92.

26. Barrios MH, Nicholson S, Bull RA, Martinello M, Rawlinson W, Mina M, et al. Comparative Longitudinal Serological Study of Anti-SARS-CoV-2 Antibody Profiles in People with COVID-19. Microorganisms. 2023 Aug;11(8):1985.

27. Renard F, Scohy A, Van der Heyden J, Peeters I, Dequeker S, Vandael E, et al. Establishing an ad hoc COVID-19 mortality surveillance during the first epidemic wave in Belgium, 1 March to 21 June 2020. Euro Surveill Bull Eur Sur Mal Transm Eur Commun Dis Bull. 2021 Dec;26(48):2001402.

28. Vernemmen C, Ekelson R, Jurčević J, Nganda S, Scohy A, Dequeker S, et al. COVID-19 Mortality Statistics: A Comparative Study of Epidemiological Surveillance Data and Death Certificates in 2020 in Belgium. Quetelet J. 2023;11(1):1–39.

29. Van Goethem N, Vilain A, Wyndham-Thomas C, Deblonde J, Bossuyt N, Lernout T, et al. Rapid establishment of a national surveillance of COVID-19 hospitalizations in Belgium. Arch Public Health Arch Belg Sante Publique. 2020 Nov 18;78(1):121.

30. Abrams S, Aerts M, Molenberghs G, Hens N. Parametric overdispersed frailty models for current status data. Biometrics. 2017 Dec;73(4):1388–400.

31. Meyer MJ, Yan S, Schlageter S, Kraemer JD, Rosenberg ES, Stoto MA. Adjusting COVID-19 Seroprevalence Survey Results to Account for Test Sensitivity and Specificity. Am J Epidemiol. 2022 Mar 24;191(4):681–8.

32. Neuhauser H, Rosario AS, Butschalowsky H, Haller S, Hoebel J, Michel J, et al. Nationally representative results on SARS-CoV-2 seroprevalence and testing in Germany at the end of 2020. Sci Rep. 2022 Nov 14;12:19492.

33. Harritshøj LH, Gybel-Brask M, Afzal S, Kamstrup PR, Jørgensen CS, Thomsen MK, et al. Comparison of 16 Serological SARS-CoV-2 Immunoassays in 16 Clinical Laboratories. J Clin Microbiol. 2021 Apr 20;59(5):10.1128/jcm.02596-20.

34. Meyer B, Torriani G, Yerly S, Mazza L, Calame A, Arm-Vernez I, et al. Validation of a commercially available SARS-CoV-2 serological immunoassay. Clin Microbiol Infect. 2020 Oct 1;26(10):1386– 94.

35. Scheiblauer H, Nübling CM, Wolf T, Khodamoradi Y, Bellinghausen C, Sonntagbauer M, et al. Antibody response to SARS-CoV-2 for more than one year − kinetics and persistence of detection are predominantly determined by avidity progression and test design. J Clin Virol. 2022 Jan 1;146:105052.

36. Lassaunière R, Frische A, Harboe ZB, Nielsen ACY, Fomsgaard A, Krogfelt KA, et al. Evaluation of nine commercial SARS-CoV-2 immunoassays [Internet]. medRxiv; 2020 [cited 2024 Mar 4]. p. 2020.04.09.20056325. Available from: https://www.medrxiv.org/content/10.1101/2020.04.09.20056325v1

37. GeurtsvanKessel CH, Okba NMA, Igloi Z, Bogers S, Embregts CWE, Laksono BM, et al. An evaluation of COVID-19 serological assays informs future diagnostics and exposure assessment. Nat Commun. 2020 Jul 6;11(1):3436.

38. Nyagwange J, Kutima B, Mwai K, Karanja HK, Gitonga JN, Mugo D, et al. Comparative performance of WANTAI ELISA for total immunoglobulin to receptor binding protein and an ELISA for IgG to spike protein in detecting SARS-CoV-2 antibodies in Kenyan populations. J Clin Virol. 2022 Jan;146:105061.

39. License and Citation – NIMBLE [Internet]. [cited 2021 Jun 18]. Available from: https://r-nimble.org/license-and-citation

40. Anda EE, Braaten T, Borch KB, Nøst TH, Chen SLF, Lukic M, et al. Seroprevalence of antibodies against SARS-CoV-2 in the adult population during the pre-vaccination period, Norway, winter 2020/21. Euro Surveill Bull Eur Sur Mal Transm Eur Commun Dis Bull. 2022 Mar;27(13):2100376.

41. Espenhain L, Tribler S, Sværke Jørgensen C, Holm Hansen C, Wolff Sönksen U, Ethelberg S. Prevalence of SARS-CoV-2 antibodies in Denmark: nationwide, population-based seroepidemiological study. Eur J Epidemiol. 2021 Jul 1;36(7):715–25.

42. Gudbjartsson DF, Norddahl GL, Melsted P, Gunnarsdottir K, Holm H, Eythorsson E, et al. Humoral Immune Response to SARS-CoV-2 in Iceland. N Engl J Med. 2020 Oct 29;383(18):1724–34.

43. Pérez-Gómez B, Pastor-Barriuso R, Fernández-de-Larrea N, Hernán MA, Pérez-Olmeda M, Oteo- Iglesias J, et al. SARS-CoV-2 Infection During the First and Second Pandemic Waves in Spain: the ENE–COVID Study. Am J Public Health. 2023 May;113(5):533–44.

44. Vos ERA, van Hagen CCE, Wong D, Smits G, Kuijer M, Wijmenga-Monsuur AJ, et al. SARS-CoV-2 Seroprevalence Trends in the Netherlands in the Variant of Concern Era: Input for Future Response. Influenza Other Respir Viruses. 2024 Jun 4;18(6):e13312.

45. Coronavirus (COVID-19) Infection Survey - Office for National Statistics [Internet]. [cited 2024 Sep 17]. Available from: https://www.ons.gov.uk/peoplepopulationandcommunity/healthandsocialcare/conditionsanddiseases/articles/coronaviruscovid19infectionsinthecommunityinengland/antibodydatafortheukjanuary2021

46. Naesens R, Mertes H, Clukers J, Herzog S, Brands C, Vets P, et al. SARS-CoV-2 seroprevalence survey among health care providers in a Belgian public multiple-site hospital. Epidemiol Infect. 2021 Jan;149:e172.

47. Wauthier L, Delefortrie Q, Eppe N, Vankerkhoven P, Wolff E, Dekeyser M, et al. SARS-CoV-2 seroprevalence in high-risk health care workers in a Belgian general hospital: evolution from the first wave to the second. Acta Clin Belg. 2022 Dec;77(6):906–14.

48. Steensels D, Oris E, Coninx L, Nuyens D, Delforge ML, Vermeersch P, et al. Hospital-Wide SARS- CoV-2 Antibody Screening in 3056 Staff in a Tertiary Center in Belgium. JAMA. 2020 Jul 14;324(2):195–7.

49. Mariën J, Ceulemans A, Bakokimi D, Lammens C, Ieven M, Heytens S, et al. Prospective SARS- CoV-2 cohort study among primary health care providers during the second COVID-19 wave in Flanders, Belgium. Fam Pract. 2022 Feb 1;39(1):92–8.

50. De Rop L, Vercruysse H, Alenus U, Brusselmans J, Callens S, Claeys M, et al. SARS-CoV-2 Seropositivity in Nursing Home Staff and Residents during the First SARS-CoV-2 Wave in Flanders, Belgium. Viruses. 2024 Sep 14;16(9):1461.

51. Janssens H, Heytens S, Meyers E, De Schepper E, De Sutter A, Devleesschauwer B, et al. Pre- vaccination SARS-CoV-2 seroprevalence among staff and residents of nursing homes in Flanders (Belgium) in fall 2020. Epidemiol Infect. 2022 Mar 2;150:1–25.

52. Peckeu-Abboud L, van Kleef E, Smekens T, Latour K, Dequeker S, Panis LI, et al. Factors influencing SARS-CoV-2 infection rate in Belgian nursing home residents during the first wave of COVID-19 pandemic. Epidemiol Infect. 2022 Jan;150:e72.

53. Janssens H, Heytens S, Meyers E, Devleesschauwer B, Cools P, Geens T. Exploratory study of risk factors related to SARS-CoV-2 prevalence in nursing homes in Flanders (Belgium) during the first wave of the COVID-19 pandemic. PloS One. 2023;18(10):e0292596.

54. Boey L, Roelants M, Merckx J, Hens N, Desombere I, Duysburgh E, et al. Age-dependent seroprevalence of SARS-CoV-2 antibodies in school-aged children from areas with low and high community transmission. Eur J Pediatr. 2022 Feb 1;181(2):571–8.

55. Cremer K, Frère J, Chatzis O, Mendonca RD, Kabamba B, Renard F, et al. SARS-CoV-2 Antibody Seroprevalence in Children and Workers from Belgian French-Speaking Primary Schools. Health (N Y). 2023 Sep 4;15(9):917–37.

56. Dethioux L, Dauby N, Montesinos I, Rebuffat E, Hainaut M. SARS-CoV-2 seroprevalence in children and their family members, July–October 2020, Brussels. Eur J Pediatr. 2022 Mar 1;181(3):1009–16.

57. Levin AT, Hanage WP, Owusu-Boaitey N, Cochran KB, Walsh SP, Meyerowitz-Katz G. Assessing the age specificity of infection fatality rates for COVID-19: systematic review, meta-analysis, and public policy implications. Eur J Epidemiol. 2020 Dec 1;35(12):1123–38.

58. Variation in the COVID-19 infection–fatality ratio by age, time, and geography during the pre- vaccine era: a systematic analysis. The Lancet. 2022 Apr 16;399(10334):1469–88.

59. Cheng C, Zhou H, Weiss JC, Lipton ZC. Unpacking the Drop in COVID-19 Case Fatality Rates: A Study of National and Florida Line-Level Data. AMIA Annu Symp Proc. 2022 Feb 21; 2021:285.

60. de Boer PT, van de Kassteele J, Vos ERA, van Asten L, Dongelmans DA, van Gageldonk-Lafeber AB, et al. Age-specific severity of severe acute respiratory syndrome coronavirus 2 in February 2020 to June 2021 in the Netherlands. Influenza Other Respir Viruses. 2023;17(8):e13174.

61. Taccone FS, Goethem NV, Pauw RD, Wittebole X, Blot K, Oyen HV, et al. The role of organizational characteristics on the outcome of COVID-19 patients admitted to the ICU in Belgium. Lancet Reg Health – Eur [Internet]. 2021 Mar 1 [cited 2021 May 11];2. Available from: https://www.thelancet.com/journals/lanepe/article/PIIS2666-7762(20)30019-3/abstract

62. Aanzienlijke oversterfte tijdens de hittegolf van augustus 2020 [Internet]. sciensano.be. [cited 2025 Mar 21]. Available from: https://www.sciensano.be/nl/pershoek/aanzienlijke-oversterfte-tijdens-de-hittegolf-van-augustus-2020

63. Molenberghs G, Faes C, Verbeeck J, Deboosere P, Abrams S, Willem L, et al. COVID-19 mortality, excess mortality, deaths per million and infection fatality ratio, Belgium, 9 March 2020 to 28 June 2020. Eurosurveillance. 2022 Feb 17;27(7):2002060.

64. Abrams S, Wambua J, Santermans E, Willem L, Kuylen E, Coletti P, et al. Modelling the early phase of the Belgian COVID-19 epidemic using a stochastic compartmental model and studying its implied future trajectories. Epidemics. 2021 Jun;35:100449.

65. García-García D, Vigo MI, Fonfría ES, Herrador Z, Navarro M, Bordehore C. Retrospective methodology to estimate daily infections from deaths (REMEDID) in COVID-19: the Spain case study. Sci Rep. 2021 May 28;11(1):11274.

66. Shioda K, Lau MSY, Kraay ANM, Nelson KN, Siegler AJ, Sullivan PS, et al. Estimating the Cumulative Incidence of SARS-CoV-2 Infection and the Infection Fatality Ratio in Light of Waning Antibodies. Epidemiology. 2021 Jul;32(4):518.

67. Takahashi S, Peluso MJ, Hakim J, Turcios K, Janson O, Routledge I, et al. SARS-CoV-2 Serology Across Scales: A Framework for Unbiased Estimation of Cumulative Incidence Incorporating Antibody Kinetics and Epidemic Recency. Am J Epidemiol. 2023 Sep 1;192(9):1562–75.

68. Fox T, Geppert J, Dinnes J, Scandrett K, Bigio J, Sulis G, et al. Antibody tests for identification of current and past infection with SARS-CoV-2. Cochrane Database Syst Rev [Internet]. 2022 [cited 2024 Jun 2];(11). Available from: https://www.cochranelibrary.com/cdsr/doi/10.1002/14651858.CD013652.pub2/full

69. Sullivan PS, Siegler AJ, Shioda K, Hall EW, Bradley H, Sanchez T, et al. Severe Acute Respiratory Syndrome Coronavirus 2 Cumulative Incidence, United States, August 2020–December 2020. Clin Infect Dis. 2022 Apr 1;74(7):1141–50.

70. Lamba K, Bradley H, Shioda K, Sullivan PS, Luisi N, Hall EW, et al. SARS-CoV-2 Cumulative Incidence and Period Seroprevalence: Results From a Statewide Population-Based Serosurvey in California. Open Forum Infect Dis. 2021 Aug 1;8(8):ofab379.

71. Buss LF, Prete CA, Abrahim CMM, Mendrone A, Salomon T, de Almeida-Neto C, et al. Three- quarters attack rate of SARS-CoV-2 in the Brazilian Amazon during a largely unmitigated epidemic. Science. 2021 Jan 15;371(6526):288–92.

72. Chen S, Flegg JA, White LJ, Aguas R. Levels of SARS-CoV-2 population exposure are considerably higher than suggested by seroprevalence surveys. PLOS Comput Biol. 2021 Sep 20;17(9):e1009436.

73. Duong S, Burtniak J, Gretchen A, Mai A, Klassen P, Wei Y, et al. Riding high: seroprevalence of SARS-CoV-2 after 4 pandemic waves in Manitoba, Canada, April 2020–February 2022. BMC Public Health. 2023 Dec 5;23(1):2420.

74. Brazeau NF, Verity R, Jenks S, Fu H, Whittaker C, Winskill P, et al. Estimating the COVID-19 infection fatality ratio accounting for seroreversion using statistical modelling. Commun Med. 2022 May 19;2(1):1–13.

75. Petrie JG, Eisenberg MC, Lauring AS, Gilbert J, Harrison SM, DeJonge PM, et al. The variant- specific burden of SARS-CoV-2 in Michigan: March 2020 through November 2021. J Med Virol. 2022;94(11):5251–9.

76. Blaizot S, Herzog SA, Abrams S, Theeten H, Litzroth A, Hens N. Sample size calculation for estimating key epidemiological parameters using serological data and mathematical modelling. BMC Med Res Methodol. 2019 Mar 7;19(1):51.

